# ADSMoE: A mixture-of-experts-based transformer autoencoder for discovering multimodal Alzheimer’s disease subtypes

**DOI:** 10.64898/2026.11.16.26368224

**Authors:** Weimin Meng, Jinqian Pan, Meichuan Yin, Pei Zhuang, Kuang Gong, Mingquan Lin, Jie Xu, Matthew Burns, Yonghui Wu, the Alzheimer’s Disease Neuroimaging Initiative, Rui Yin

**Author notes:** Correspondence to Rui Yin, PhD Department of Health Outcomes & Biomedical Informatics University of Florida Gainesville, Florida, USA (R.Y.). This study used data from the Alzheimer’s Disease Neuroimaging Initiative (ADNI) database (adni.loni.usc.edu).

## Abstract

Alzheimer’s disease (AD) exhibits substantial biological and clinical heterogeneity, posing critical challenges for advancing precision medicine. Existing data-driven approaches often rely on modality-specific clustering or predefined biomarker trajectories, limiting their ability to discover disease subtypes from incomplete, heterogeneous real-world data. In this study, we introduce ADSMoE, a mixture-of-experts (MoE)-based multimodal representation learning framework that jointly identifies AD subtypes as latent disease signatures while accommodating incomplete modality combinations. ADSMoE integrates a Fleximodal MoE encoder that accommodates arbitrary combinations of available modalities with a novel subtype-specific MoE decoder to capture complementary disease characteristics through expert specialization. Using data from 5,422 participants across three independent cohorts, we trained ADSMoE on the ADNI cohort and externally validated the identified subtypes in NACC and UKB without model retraining. The proposed ADSMoE identified four AD subtypes with distinct neuroimaging patterns, genetic and proteomic associations, and clinical characteristics. Benchmarking analyses demonstrated robust subtype discovery and strong biological and clinical relevance, while external validation revealed consistent subtype-associated characteristics across independent cohorts. ADSMoE advances AD subtyping by uncovering biologically and clinically meaningful AD heterogeneity from incomplete multimodal data, providing a scalable framework for disease stratification, biomarker discovery, and future precision medicine applications in AD.

## Introduction

Alzheimer’s disease (AD) is a biologically and clinically heterogeneous neurodegenerative disorder with substantial variability in pathology and progression. As of 2025, AD affects approximately 7 million individuals in the United States, a number projected to nearly double to 13.85 million by 2060^1^. The rapidly increasing prevalence of AD presents significant challenges not only for affected individuals and their families but also for healthcare systems and society. Disease heterogeneity manifests across cognitive impairment, neurodegenerative patterns, biomarker profiles, genetic risk factors, and rates of clinical progression^2^, highlighting the need to identify biologically and clinically meaningful AD subtypes. Recent advances in multimodal features, such as neuroimaging and fluid–based biomarkers, have provided important opportunities to characterize AD heterogeneity^3^. For example, positron emission tomography (PET) measures of amyloid–β (Aβ) and tau^4,5^, while magnetic resonance imaging (MRI)–based measures of structural and functional brain changes capture complementary aspects of AD pathology and neurodegeneration^6^. Blood- and cerebrospinal fluid (CSF)-based biomarkers^7,8^ further reflect molecular alterations associated with AD. Genetic studies have identified susceptibility variants in genes such as *APOE* and *TREM2*^9^ and pathogenic mutations in *APP*, *PSEN1*, and *PSEN2* linked to early-onset AD^10,11^. Importantly, these biomarkers are not independent but interact across molecular, structural, and functional levels. Prior studies have reported that neuroimaging abnormalities are associated with genetic risk variants^12,13^, and CSF biomarkers closely correspond to Aβ and tau pathology detected by neuroimaging^14^. Moreover, growing evidence indicates that blood- and CSF-based biomarkers are influenced by both *APOE* genotype and broader polygenic risk profiles^15,16^. These complementary and interconnected biomarkers provide a strong biological foundation for multimodal characterization of AD heterogeneity.

Despite these advances, effectively integrating multimodal biomarkers into large-scale, real-world studies and clinical practice remains challenging. In longitudinal cohort and routine clinical settings, multimodal data are often incompletely acquired, as neuroimaging, fluid biomarkers, genetic profiling, and detailed clinical assessments are collected at different time points, under varying protocols, or only in selected patient subsets^17–20^. Consequently, incomplete modality availability is the rule rather than the exception, leading to heterogeneous observations across individuals and constraining the characterization of AD heterogeneity and disease progression in real-world populations^5,21,22^. Recent developments in artificial intelligence (AI) approaches have improved our understanding of AD heterogeneity but remain limited in modeling AD subtypes as latent signatures jointly learned from heterogeneous multimodal data^23–25^. Current methods generally fall into three categories. First, progression-based frameworks, such as Subtype and Stage Inference (SuStaIn) and its extensions, disentangle disease stage from phenotypic variability by modeling biomarker evolution over time^20,26–28^, but they rely on predefined progression trajectories and typically operate on a limited subset of consistently acquired biomarkers. Second, modality-centered subtype discovery approaches, such as GeneSGAN^29^, incorporate genetic information to improve biological interpretation but generally anchor subtype identification to a dominant modality, most commonly neuroimaging, while treating other modalities as auxiliary information. Consequently, the resulting subtypes represent modality-specific phenotypes rather than shared multimodal disease signatures. Third, recent multimodal deep learning frameworks based on autoencoders and transformers have demonstrated powerful representation learning across neuroimaging, genetic, molecular, and clinical data^30–38^. However, these models are primarily designed for prediction or classification tasks, with subtype identification performed only as a downstream analysis, and generally assume a fixed set of input modalities during training. These assumptions limit robustness to irregular modality availability and restrict applicability to large-scale, real-world cohorts, where missing or partially observed modalities are common. Overall, existing approaches either learn modality-specific representations or perform multimodal prediction without explicitly modeling disease subtypes as latent multimodal signatures emerging from heterogeneous and incomplete real-world observations.

Given the heterogeneous and incomplete nature of real-world AD data, the Mixture–of–Experts (MoE) framework provides a flexible, adaptive strategy for multimodal representation learning^39,40^. By allowing different experts to specialize in complementary multimodal patterns, MoE naturally accommodates irregular modality availability and preserves complementary information across modalities. Previous studies have demonstrated the effectiveness of MoE architectures for multimodal representation learning, including multimodal embedding learning^41,42^, multi–omics integration^43^, brain foundation models^44^, and learning under missing modalities^41,45^. More recently, advanced MoE-based transformer architectures have further improved training stability and expert specialization^46^. In the context of multimodal AD data, we hypothesize that routed experts can capture disease characteristics and learn subtype-specific patterns, enabling biologically and clinically meaningful disease heterogeneity to emerge through multimodal representation learning.

In this work, we present ADSMoE (**A**lzheimer’s **D**isease **S**ubtyping via **M**ixture-**o**f-**E**xperts), an MoE-based multimodal representation learning framework for discovering AD subtypes from heterogeneous real-world data. Unlike existing approaches, ADSMoE integrates incomplete and irregular multimodal data through a flexible-modal MoE encoder and introduces a novel subtype-specific MoE decoder that learns disease patterns through reconstruction. Using ADSMoE, we identified four biologically and clinically distinct AD subtypes characterized by significant differences in neuroanatomical patterns, molecular signatures, genetic variation and clinical characteristics. These four subtypes were discovered by training ADSMoE model on the ADNI dataset and validated in independent NACC and UKB cohorts, with consistent multimodal characteristics. These findings establish a generalizable framework for discovering reproducible AD subtypes from incomplete multimodal data and demonstrate the potential of an MoE-based transformer autoencoder to capture AD heterogeneity. It provides a scalable foundation for precision diagnosis, biomarker discovery, and subtype-guided therapeutic development in Alzheimer’s disease.

## Results

### Overview of the ADSMoE framework

ADSMoE is a novel MoE–based transformer autoencoder designed to identify AD subtypes using heterogeneous multimodal data, integrating imaging, genetics, clinical, and protein data from three large cohorts (i.e., ADNI, NACC, and UKB). As shown in **Figure 1**, ADSMoE comprises a modality-flexible transformer encoder (termed the Fleximodal^47^ encoder) that can accept an arbitrary combination of input modalities, and an MoE-based transformer decoder that learns subtype-specific AD representations (**Figure 1A**). The encoder generates unified latent representations, which are used to initialize subtype structure through the expectation-maximization (EM) algorithm^48^ and calculate decoder gate weights. These weights subsequently guide expert specialization within the decoder, allowing subtype-specific experts to progressively capture distinct multimodal disease patterns^40^. The decoder reconstructs latent representations and learns subtype patterns. Unlike conventional clustering approaches that assign patients solely according to feature similarity, ADSMoE learns subtype assignments jointly with multimodal representation learning through expert routing (**Figure 1B**). Potential AD subtypes, which are patient groups with different disease-related characteristics, are routed to subtype-specific experts, while severity is modeled separately to avoid being driven dominantly by disease stage. The model is optimized in three sequential phases (**Figure 1C**). First, normative modeling^49,50^ is introduced to quantify individual deviations from healthy reference while reducing the effects of disease-unrelated confounders, converting each modality into residuals. Second, the Fleximodal encoder is trained to learn unified multimodal representations. Finally, the model jointly optimizes all components (e.g., the encoder, decoder, and routing network) to specialize each expert in a subtype-specific disease pattern. After convergence, each AD patient is assigned to the subtype with the highest routing probability (**Figure 1D**), followed by post-hoc analyses that characterize each subtype’s biological and clinical relevance. Overall, ADSMoE provides a flexible and unified framework for discovering AD subtypes from heterogeneous multimodal data and characterizing their distinct biological signatures across independent cohorts.

**Figure 1.**
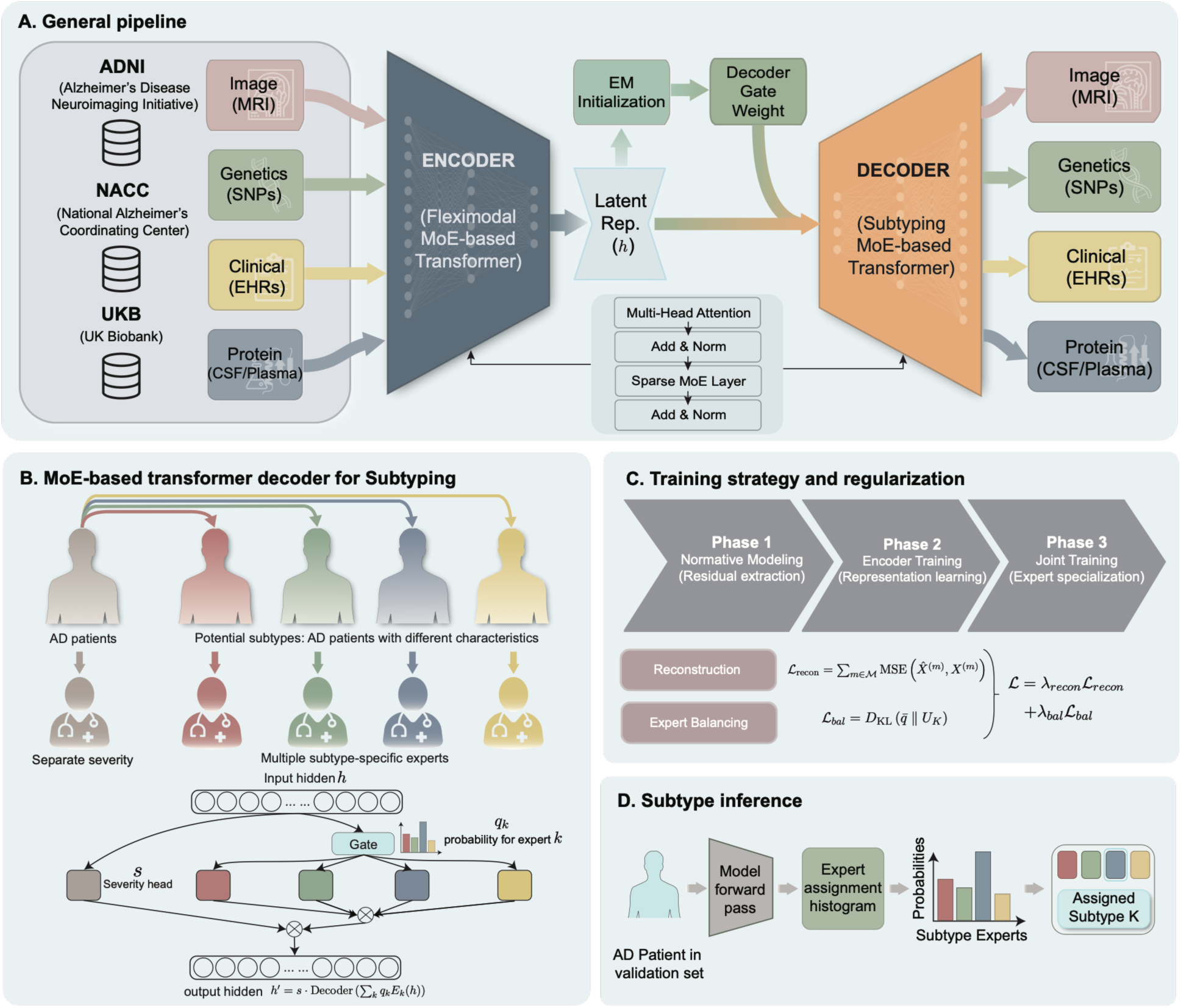
The overview of the ADSMoE framework for multimodal representation learning and AD subtyping. (**A)** Overall workflow of ADSMoE, integrating multimodal data from multiple cohorts, using a Fleximodal MoE-based transformer autoencoder for unified representation learning and subtyping. (**B**) Architecture of the MoE-based transformer decoder, comprising multiple subtype-specific experts for learning distinct multimodal disease patterns through expert routing. (**C**) Three-stage training strategy, including normative modeling, encoder training, and joint training following EM-based initialization, with reconstruction and expert-balancing losses to promote multimodal representation learning and subtype-specific expert specialization. (**D**) Subtype inference based on expert routing probabilities, assigning each patient to the AD subtype with the highest routing probability.

### Study cohorts and characteristics

In this study, we collected AD cohorts from three large data resources: the Alzheimer’s Disease Neuroimaging Initiative (ADNI)^51^, the National Alzheimer’s Coordinating Center (NACC)^52^, and the UK Biobank (UKB)^53^, which collectively provide complementary multimodal data, including clinical, genetic, protein, and image data. For ADNI, we included all participants. For NACC and UKB, we included cases with at least two modalities. We ended up with 2,035, 1,635, and 1,752 participants from ADNI, NACC, and UKB, respectively. Although the NACC and UKB have substantially more eligible participants with AD, we randomly selected 876 individuals, the same as ADNI. As shown in **Table 1**, participants with AD were predominantly older adults across all cohorts, with most individuals aged between 65 and 84 years (74.9% - 93.3%), and a similar mean age of approximately 75 years. Female participants accounted for a lower proportion in ADNI (39.3%) but a slightly higher proportion in the NACC and UKB cohorts (54.3% and 55.1%). APOE ε4 carriage was common across cohorts. The proportion of individuals carrying two copies of APOE ε4 alleles varied substantially, ranging from 7.8% in NACC to 22.4% in UKB. The three cohorts also differed in multimodal data availability, providing complementary evaluation settings for ADSMoE. ADNI involved all selected modalities with varying completeness, NACC mainly contributed to clinical and imaging data, whereas UKB provided comprehensive genetic and protein biomarker data but relatively limited MRI measurements. These heterogeneous modality combinations reflect the incomplete and irregular data commonly encountered in real-world AD studies and provide a suitable benchmark for evaluating the robustness and generalizability of ADSMoE. Additional cohort characteristics are provided in the **Supplementary Table S1**.

**Table 1.** Baseline characteristics of participants with AD across ADNI, NACC, and UKB. Demographic characteristics, APOE ε4 status, and modality availability are summarized. Data are presented as n (%) for categorical variables and mean (standard deviation [SD]) for continuous variables.

| **Cohort** | **ADNI**<br>AD<br>(N = 876) | **NACC**<br>AD<br>(N = 876) | **UKB**<br>AD<br>(N = 876) |
|---|---|---|---|
| **Age** |  |  |  |
| <65 | 91 (10.4%) | 123 (14.0%) | 58 (6.6%) |
| 65-84 | 656 (74.9%) | 675 (77.1%) | 817 (93.3%) |
| >=85 | 58 (6.6%) | 78 (8.9%) | 1 (0.1%) |
| Unknown | 71 (8.1%) | 0 (0.0%) | 0 (0.0%) |
| Mean (SD) | 74.5 (7.4) | 73.8 (8.5) | 74.7 (6.2) |
| **Sex** |  |  |  |
| Male | 461 (52.6%) | 400 (45.7%) | 393 (44.9%) |
| Female | 344 (39.3%) | 476 (54.3%) | 483 (55.1%) |
| Unknown | 71 (8.1%) | 0 (0.0%) | 0 (0.0%) |
| **Race** |  |  |  |
| White | 748 (85.4%) | 688 (78.5%) | 850 (97.0%) |
| Black or African American | 33 (3.8%) | 144 (16.4%) | 13 (1.5%) |
| American Indian or Alaska Native | 0 (0.0%) | 8 (0.9%) | 0 (0.0%) |
| Native Hawaiian or Pacific Islander | 0 (0.0%) | 0 (0.0%) | 0 (0.0%) |
| Asian | 16 (1.8%) | 25 (2.9%) | 2 (0.2%) |
| Multiracial | 7 (0.8%) | 5 (0.6%) | 4 (0.5%) |
| Unknown or ambiguous | 72 (8.2%) | 6 (0.7%) | 7 (0.8%) |
| **Hispanic ethnicity** |  |  |  |
| No | 777 (88.7%) | 814 (92.9%) | - |
| Yes | 24 (2.7%) | 59 (6.7%) | - |
| Missing/Unknown | 75 (8.6%) | 3 (0.3%) | - |
| **APOE Biomarkers** |  |  |  |
| No APOE ε4 allele | 268 (30.6%) | 355 (40.5%) | 281 (32.1%) |
| 1 copy of APOE ε4 allele | 376 (42.9%) | 256 (29.2%) | 380 (43.4%) |
| 2 copies of APOE ε4 allele | 133 (15.2%) | 68 (7.8%) | 196 (22.4%) |
| Missing or unknown or not assessed | 99 (11.3%) | 197 (22.5%) | 19 (2.2%) |
| **Modality Availability** |  |  |  |
| Image MRI | 848 (96.8%) | 876 (100.0%) | 61 (7.0%) |
| Protein Measurements | 329 (37.6%) | 0 (0.0%) | 647 (73.9%) |
| Genetics | 734 (83.8%) | 0 (0.0%) | 876 (100.0%) |
| Clinical | 740 (84.5%) | 876 (100.0%) | 876 (100.0%) |

### Model performance

#### Benchmark evaluation

We first benchmarked several encoders to identify the optimal structure for multimodal representation learning by discriminating individuals with AD from cognitively normal (CN) participants using ADNI dataset. As shown in **Figure 2A**, FlexMoE achieved the best overall performance, with the highest results in both the validation (accuracy = 0.906, F1 = 0.890, AUC = 0.936) and the test sets (accuracy = 0.892, F1 = 0.871, AUC = 0.938), while maintaining computational efficiency (192.0 s per run). Although FuseMoE showed competitive predictive performance, it required substantially longer runtime (309.4 s per run). In contrast, ResNet and LiMoE were more computationally efficient (12.5 s and 51.1 s per run), but consistently underperformed across all evaluation metrics. As a result, we selected FlexMoE as the multimodal encoder for ADSMoE. We next evaluated subtype discovery on ADNI by comparing ADSMoE with several representative methods (**Figure 2B)**. Because ground-truth AD subtype labels are unavailable in real-world cohorts, we evaluated subtype quality using the Davies-Bouldin index (DBI)^54^ and a genetics-informed metric, i.e., the number of significantly associated single-nucleotide polymorphisms (denoted as N-Asso-SNPs)^29^. Lower DBI indicates better clustering compactness and separation, while higher N-Asso-SNPs suggests stronger genetic relevance of the identified subtypes. Additional evaluations under varying multimodal missingness, including different proportions of genetic variants, region of interest (ROI)-level MRI features^55^, clinical variables, and AD-related protein biomarkers, are presented in **Supplementary Figure S1**. Results are reported as mean ± standard deviation (SD) from five-fold cross-validation. Across all comparisons, ADSMoE consistently achieved the strongest genetic association signals (N–Asso–SNPs = 189 ± 16) while maintaining a low DBI (4.24 ± 0.74), indicating genetically related and well-separated subtype structures. In comparison, GeneSGAN identified substantially fewer associated SNPs (100 ± 9) and exhibited poorer clustering quality (DBI = 22.57 ± 0.46). CDEC produced highly compact clusters (DBI = 0.46 ± 0.00) but yielded limited genetic associations (N–Asso–SNPs = 59 ± 18), suggesting that geometric compactness alone does not necessarily correspond to biologically meaningful subtypes. EDESC had intermediate performance (N–Asso–SNPs = 64 ± 6, DBI = 13.75 ± 0.48), whereas classical clustering methods (Kmeans and spectral clustering) performed consistently worse across metrics. Overall, ADSMoE achieved the strongest subtype-genetic associations among the compared methods while maintaining competitive clustering quality.

**Figure 2.**
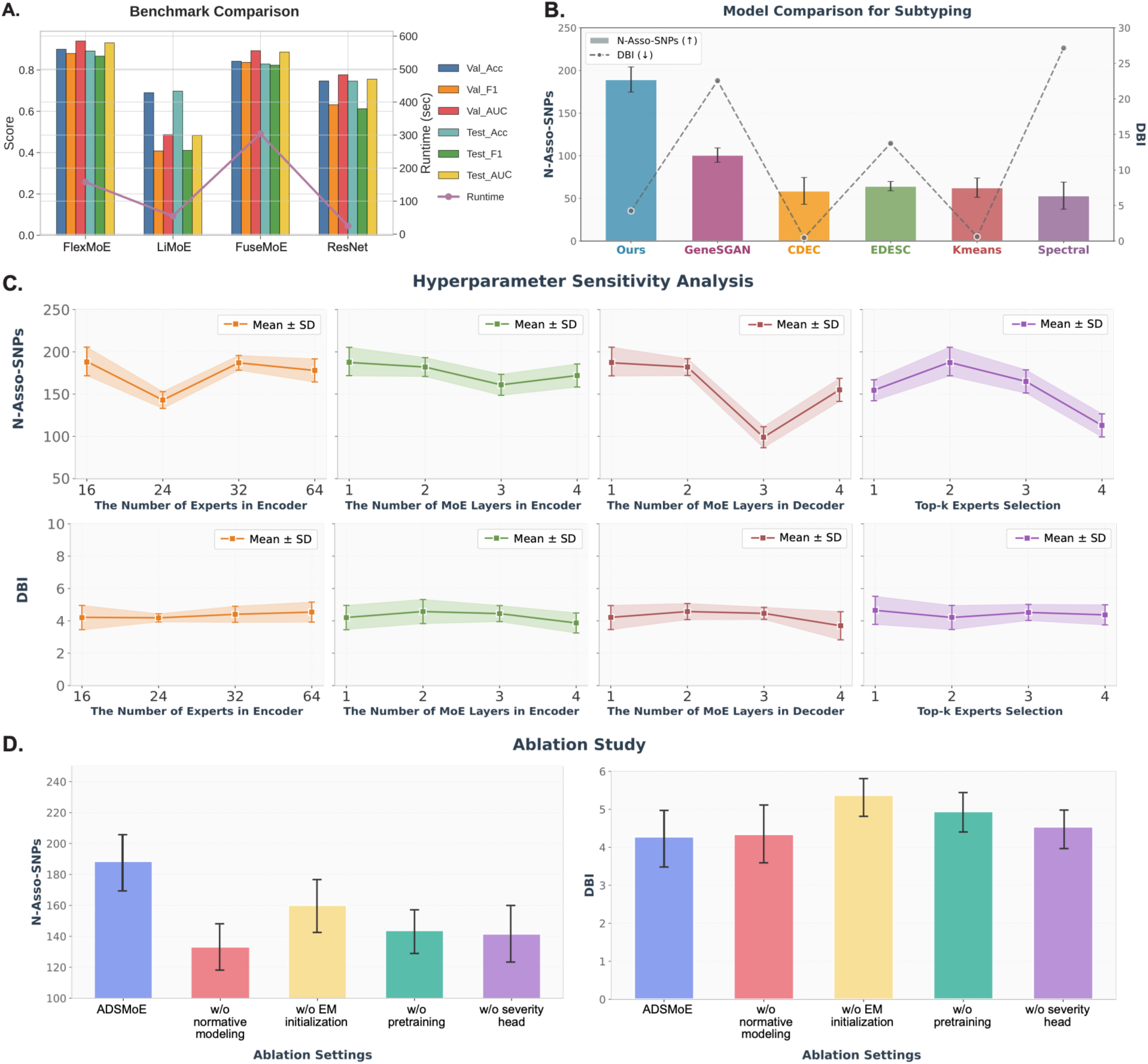
Performance evaluation, hyperparameter sensitivity, and ablation analysis of ADSMoE. (**A**) Benchmark comparison of multimodal encoders for distinguishing AD from CN participants. (**B**) Subtyping comparison between ADSMoE and several representative methods. (**C**) Hyperparameter sensitivity analysis evaluating the effects of encoder expert number, encoder and decoder MoE depth, and top-*k* expert selection. (**D**) Ablation study assessing the contributions of key components of normative modeling, EM initialization, decoder gate weight calculation, and the severity head.

#### Hyperparameter sensitivity analysis

To assess the robustness of ADSMoE, we performed a sensitivity analysis. ADSMoE demonstrated stable subtype discovery performance across different hyperparameter configurations (**Figure 2C**). Varying the number of experts from 16 to 32 in the encoder showed only modest changes in both N–Asso–SNPs and DBI, while N-Asso-SNPs decreased more when changing to 24 experts. When increasing the number of MoE layers to four in the encoder, we found it slightly improved cluster compactness (lower DBI) but did not strengthen genetic associations, suggesting diminishing returns from deeper encoder architectures. Increasing decoder depth to three layers of MoE reduced N–Asso–SNPs, whereas DBI remained lower with more decoder MoE layers, which shows a difference between biological relevance and clustering quality. Finally, varying the top-k routing strategy had little effect on DBI but influenced genetic association strength, with top-2 routing achieving the highest N-Asso-SNPs.

#### Contribution of ADSMoE modules

We further performed an ablation study to evaluate the contribution of key modules of ADSMoE. As shown in **Figure 2D**, the full model achieved the highest N–Asso–SNPs (189 ± 16) while maintaining a low DBI (4.24 ± 0.74). Removing normative modeling, which reduces confounders through residualization, produced a noticeable reduction in N–Asso–SNPs (132 ± 15) and increased DBI (4.32 ± 0.81), showing that normative modeling is critical for identifying disease-related subtype representations. In contrast, removing EM initialization resulted in the largest deterioration in clustering performance, yielding both reduced N–Asso–SNPs (159 ± 18) and the highest DBI (5.33 ± 0.47), highlighting the importance of clustering initialization. Moreover, eliminating either pretraining or the severity head reduced N-Asso-SNPs (142 ± 16 and 141 ± 19, respectively), highlighting their contributions to meaningful subtype discovery. Removing pretraining also increased DBI (4.85 ± 0.57), suggesting that pretraining improves clustering quality. Overall, these results show that each component contributes to the overall performance of ADSMoE.

### Characterization of ADSMoE-derived AD subtypes

#### Latent representation and modality robustness

Using ADSMoE, we identified four multimodal subtypes in the ADNI dataset. We visualized the latent embeddings generated by the FlexMoE encoder using principal component analysis (PCA)^56^, t-distributed stochastic neighbor embedding (t-SNE)^57^, and uniform manifold approximation and projection (UMAP)^58^. As shown in **Figure 3A**, the four identified subtypes formed distinguishable clusters using PCA, although partial overlap was observed between neighboring subtypes, reflecting the continuous nature of AD heterogeneity. Similar subtype separation was consistently observed using t-SNE and UMAP (**Supplementary Figure S2**), supporting the robustness of the learned latent representations. We further investigated whether subtypes were associated with specific modality combinations by summarizing subtype distributions across different patterns of available modalities (**Figure 3B**), where, P, C, G, and I denote protein, clinical, genetic, and image modalities, respectively. All four subtypes were represented across multiple modality combinations rather than being confined to a single data configuration. PCGI was the most common modality combination in S1 (36.7%), S2 (33.9%), and S4 (43.8%), whereas CGI was the most common in S3 (40.9%). These findings indicate that ADSMoE learns subtype-specific disease representations that are robust to heterogeneous multimodal availability, rather than simply reflecting missing-data patterns or modality composition.

**Figure 3.**
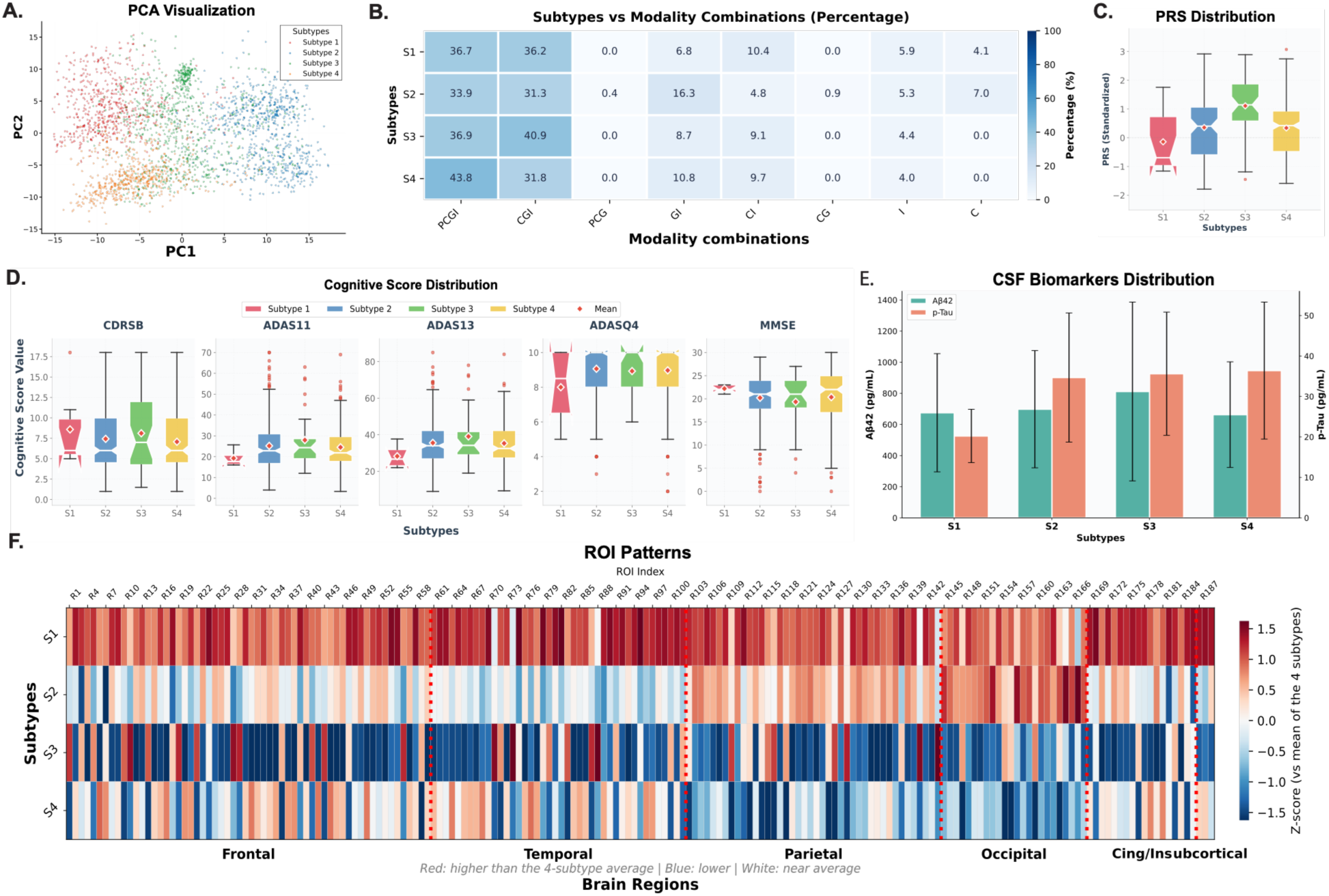
Multimodal characterization of identified AD subtypes. (**A**) Visualization of the learned latent representations of AD subtypes using PCA. (**B**) Distribution of subtypes across different modality combinations. (**C**) Comparison of PRS distributions across the four subtypes. (**D**) Cognitive performance across subtypes measured by several cognitive assessments. (**E**) CSF biomarker profiles (Aβ42 and p-tau) across subtypes. (**F**) Regional neuroimaging patterns across subtypes, summarized by ROI-level z-score distributions relative to the four-subtype mean for each ROI in major brain regions.

#### Multimodal characterization of identified subtypes

To characterize the biological and clinical profiles of the identified AD subtypes, we compared Polygenic Risk Score (PRS)^59^, cognitive performance, cerebrospinal fluid (CSF) biomarkers, and ROI patterns (**Figure 3C-F**). Overall, the four identified subtypes exhibited distinct genetic, clinical, and neurobiological profiles, supporting the biological relevance of the subtype discovery. As shown in **Figure 3C**, genetic risk differed across subtypes, with S3 showing the highest PRS overall (1.324 ± 0.961), whereas S1 exhibited the lowest (−0.155 ± 0.977) genetic risk. Consistent with this pattern, S3 also demonstrated the greatest cognitive impairment (**Figure 3D**), characterized by higher Alzheimer’s disease assessment scales (ADAS)^60^ and lower mini-mental state examination (MMSE) scores. In contrast, S1 showed the least impaired performance, while Subtype 2 (S2) and Subtype 4 (S4) displayed intermediate levels of cognitive dysfunction. Subtype-specific differences were also observed in CSF biomarkers and neuroimaging features. As shown in **Figure 3E**, p-Tau concentrations were higher in S2–S4 than in S1, whereas Aβ42 levels were highest in S3. Analysis of imaging-derived regional ROI z-score patterns across major brain regions further revealed distinct neuroanatomical signatures across subtypes (**Figure 3F**). S3 showed widespread reductions in ROI z-scores across multiple cortical regions, whereas S1 exhibited values higher than the other subtypes on average. In contrast, S2 displayed relatively lower z-scores in frontal and temporal regions but higher values in occipital regions, while S4 showed more pronounced reductions in parietal and occipital regions. In addition, demographic characteristics also differed among subtypes (**Supplementary Table S2**). S4 contained relatively younger participants (71.9 ± 7.5), whereas S2 included older participants on average (76.4 ± 6.8). S3 had more males (54.4%), while S4 (50.6% male) showed a more balanced sex distribution. Additional differences were observed in racial composition and educational attainment, indicating that the identified subtypes capture heterogeneity extending beyond molecular and neuroimaging characteristics.

### Biological and clinical signatures of AD subtypes

#### Subtype-specific structural MRI patterns

We assessed subtype-specific structural MRI patterns by ROI–level comparisons of cortical volume for each identified subtype, using Welch’s two–sample t–tests (one–vs–rest). The resulting subtype-specific ROI t-statistic maps are presented in **Figure 4A**. Overall, the four subtypes exhibited distinct spatial patterns of cortical volume variation, supporting the presence of heterogeneous neuroanatomical phenotypes. S3 demonstrated predominantly negative t–statistics across widespread cortical regions, with marked differences in the frontal, temporal, and parietal lobes, as well as in the insula, hippocampus, cingulate, and supramarginal regions, indicating broadly reduced cortical volumes compared with the remaining participants. In contrast, S1 exhibited widespread positive t-statistics across frontal, temporal, parietal, and occipital regions, suggesting relatively preserved cortical volumes. S2 also showed predominantly positive t-statistics, particularly in frontal and occipital regions, with relatively smaller differences in temporal regions. S4 displayed a more regionally heterogeneous pattern, characterized by positive t-statistics in temporal regions and negative values in portions of the frontal, parietal, and occipital cortices. These subtype-specific t-statistic maps highlight heterogeneous neuroanatomical patterns across subtypes, indicating that the identified subtypes are associated with distinct spatial profiles of cortical volume variations. Additional pairwise subtype comparisons are provided in the **Supplementary Figure S3**.

**Figure 4.**
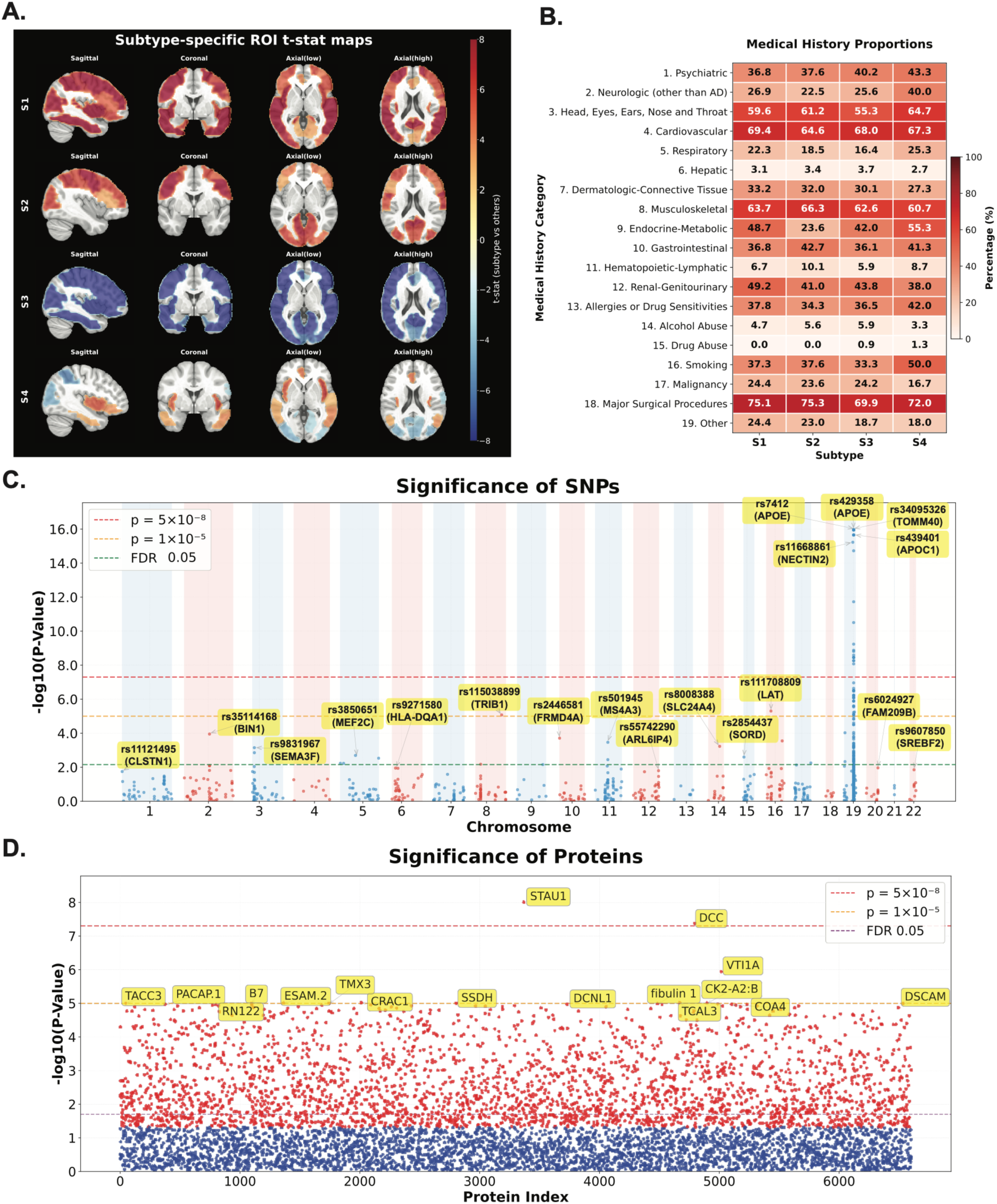
Subtype-specific imaging patterns and multimodal associations of identified AD subtypes. **(A)** Subtype-specific structural MRI signatures visualized as ROI-level t-statistic maps from one-versus-rest comparisons. (**B**) Medical history profiles across subtypes, summarized by the prevalence of major comorbidity categories. (**C**) Significantly associated SNPs identified through SNP-subtype association analysis. (**D**) Significantly associated proteins identified through protein–subtype association analysis.

#### Prevalence of medical conditions

Differences in the prevalence of medical history conditions were also observed across the four identified subtypes (**Figure 4B**). According to the results, S4 exhibited the highest prevalence of psychiatric conditions (43.3%) and smoking history (50.0%), whereas S1 showed the highest prevalence of cardiovascular disease (69.4%). In contrast, hepatic disorders, alcohol abuse, and drug abuse were uncommon across all subtypes. Other common conditions, including musculoskeletal disorders and head, eye, ear, nose, and throat conditions, were observed across subtypes but varied more modestly in prevalence. Major surgical procedures were most prevalent in S2 (75.3%) and least prevalent in S3 (69.9%). Overall, these findings demonstrate that the ADSMoE-derived subtypes are associated with clinically distinct comorbidity profiles, further supporting their clinical relevance.

#### Subtype-specific genetic associations

To investigate the genetic basis of the identified AD subtypes, we evaluated SNP-subtype associations using likelihood-ratio tests (LRTs) comparing multinomial logistic regression models fitted with and without each SNP (**Figure 4C**). The Manhattan plot revealed a pronounced enrichment of association signals on chromosome 19, where multiple variants exceed the genome-wide significance threshold. These associations were dominated by the *APOE* region, including *rs429358 and rs7412* (*APOE*), together with neighboring variants in *TOMM40* (*rs34095326*), *APOC1* (*rs439401*), and *NECTIN2* (*rs11668861*), highlighting the central role of this genomic region in distinguishing AD subtypes. Beyond chromosome 19, variants in *TRIB1* (*rs115038899*) and *LAT* (*rs111708809*) reached the suggestive significance threshold, and variants in *BIN1* (*rs35114168*), *FRMD4A* (*rs2446581*), *MS4A3* (*rs501945*), and *SLC24A4* (*rs8008388*) were significant after FDR correction. Although these associations were less significant than those observed at the APOE locus, they were distributed across multiple chromosomes, suggesting that the identified subtypes are influenced by polygenic genetic architectures rather than a single dominant susceptibility locus.

#### Subtype-specific protein associations

We further compared protein abundances across subtypes using one-way ANOVA to characterize molecular differences among the identified subtypes. The results are shown in **Figure 4D**, and we found that several proteins showed significant subtype-dependent differences. *STAU1* showed the strongest association among all measured proteins, followed by *DCC*, *VTI1A*, *CRAC1*, *TCAL3*, *CK2-A2:B*, *ESAM.2*, *TMX3*, and *TACC3*. Other significantly associated proteins are involved in diverse biological pathways, such as immune and inflammatory signaling (e.g., *RIPK2*, *CC134*), lipid metabolism (e.g., *ApoB*), and cellular stress or translational regulation (e.g., *IF4E2*). The broad functional diversity of these proteins suggests that the identified AD subtypes differ across multiple molecular pathways rather than a single biological process, further supporting their biological heterogeneity.

### Multimodal association and interpretation

#### Cross-modal associations

To investigate cross-modal relationships underlying the identified subtypes, we analyzed associations among genetic variants, protein biomarkers, neuroimaging features, and clinical variables. All features were adjusted for age and sex prior to integration to minimize potential confounding effects. The resulting network revealed extensive links across modalities (**Figure 5A**), with genetic variants exhibiting the most extensive cross-modal connectivity, particularly with structural MRI features. Multiple AD-associated variants, including those within the *APOE* locus (e.g., *APOE*, *APOC1*, and *NECTIN2*), showed widespread associations with cortical regions spanning the frontal, temporal, and parietal lobes. These findings suggest that subtype-specific genetic variation is closely linked to distributed patterns of brain structural alteration. For proteins, *STAU1* showed a positive similarity with *rs429358* (0.713) and a negative similarity with the left superior parietal MRI feature (−0.722). Clinical variables also demonstrated prominent cross-modal associations. The tremor measure was positively associated with MRI features in frontal, temporal, and parietal regions. The 18 years of education also exhibited associations with the left superior parietal MRI feature and several proteins, including *DCC*, further supporting the influence of demographic and clinical factors on subtype-specific neuroanatomical variation. The complete similarity matrix is provided in the **Supplementary Data**.

**Figure 5.**
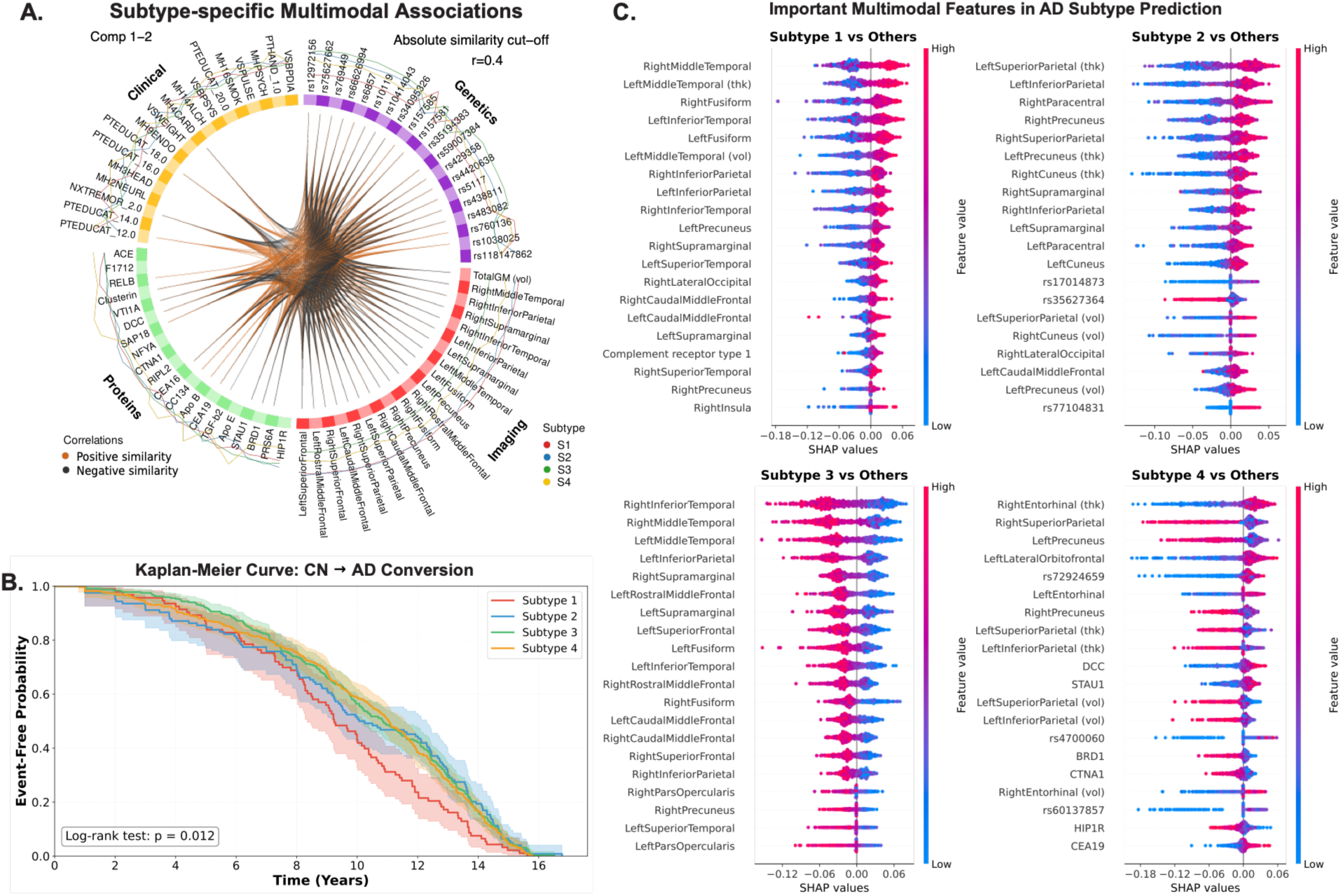
Cross-modal relationships, disease progression, and multimodal feature importance of identified AD subtypes. (**A**) Cross-modal similarity network linking genetic variants, structural MRI measures, clinical variables, and protein biomarkers across the identified subtypes. Only associations with an absolute similarity score |r| > 0.4 are shown. Edge colors indicate positive (orange) and negative (black) similarities, and nodes are grouped by modality and subtype. (**B**) Kaplan-Meier survival curves comparing time retrospectively across the four subtypes for conversion from CN to AD. Statistical differences between survival curves were assessed using a log-rank test. (**C**) Multimodal important features for one-versus-rest subtype prediction based on SHAP values. The top-ranked features for each subtype are shown, ranked by mean absolute SHAP value.

#### Survival analysis

To evaluate whether time to AD diagnosis differed across the identified subtypes, we retrospectively compared time to disease conversion (CN → AD) in each subtype using Kaplan-Meier analysis (**Figure 5B**). The four subtypes exhibited significantly different conversion trajectories (log-rank test: *p* = 0.012). During early follow-up, the survival curves largely overlapped, although S2 showed a modestly faster decline in event-free probability within the first five years. The curves began to diverge during mid-follow-up (approximately 5-7 years), with S1 demonstrating the earliest and most rapid decline, whereas S3 and S4 maintained higher event-free probabilities for a longer duration. By the end of follow-up (∼14-16 years), event-free probability approached zero across all subtypes, resulting in the convergence of survival curves. These findings indicate that the identified subtypes exhibit distinct temporal patterns of observed time to AD diagnosis intervals. Additional Kaplan-Meier analyses for CN → mild cognitive impairment (MCI)^61^ and MCI → AD transitions are provided in **Supplementary Figure S4**.

#### Multimodal feature importance

To identify the multimodal features driving subtype classification, we computed SHAP values for one–vs–rest prediction models and summarized the top-ranked features for each subtype (**Figure 5C**). Across all subtypes, MRI-derived regional measures dominated the top-ranked features, with genetic variants and CSF proteins among the top predictors for some subtypes. Each subtype exhibited a distinct combination of predictive features. S1 was primarily characterized by cortical thickness in the right and left middle temporal gyri and the right fusiform gyrus, with complement receptor type 1 also among the top-ranked features. S2 was distinguished by cortical thickness in the left superior and inferior parietal lobules and the right paracentral lobule, and variants involving *rs17014873* and *rs35627364*. For S3, all 20 top-ranked features were cortical thickness measures, led by the right inferior temporal and bilateral middle temporal regions. S4 combined right entorhinal, right superior parietal, and left precuneus cortical thickness, with the *rs72924659* variant and *DCC* and *STAU1*. Notably, several features consistently ranked among the most important predictors (e.g., left inferior parietal and right precuneus cortical thickness) across multiple subtypes. Overall, these findings suggest that subtype identification is driven by coordinated multimodal biomarker signatures.

### External validation of AD subtypes

#### Subtype characterization in the NACC cohort

We applied ADSMoE model to the independent NACC cohort without retraining or fine-tuning to evaluate the generalizability of the identified AD subtypes. Consistent with the subtype characteristics observed in ADNI, significant differences in cognitive performance of identified AD subtypes (S1-S4) were observed in the NACC cohort (**Figure 6A**). These differences were assessed using five commonly used cognitive measures including Clinical Dementia Rating – Sum of Boxes (CDR-SB) scores^62^, Montreal Cognitive Assessment (MoCA) scores^63^, delayed story recall^64^, animal fluency^65^, and Trail Making Test B^66^ completion times, which assess cognitive function and impairment similar to those evaluated in ADNI. S3 showed the greatest cognitive impairment, characterized by higher CDR-SB, lower MoCA scores, poorer delayed story recall and animal fluency, and longer Trail Making Test B completion times. In contrast, S1 and S2 exhibited better cognitive performance, whereas S4 showed intermediate impairment. MRI analyses revealed neuroanatomical patterns consistent with those in ADNI (**Figure 6B**), with S1 showing widespread positive ROI t-statistics, S3 exhibiting predominantly negative values across cortical regions, and S2 and S4 displaying more region-specific patterns. Genetic differences were also observed (**Figure 6C**), with APOE ε4 carriage varying significantly across subtypes (χ² test, *p* = 0.029), providing genetic evidence independent of the inputs used for subtype assignment. S3 exhibited the highest proportion of APOE ε4 carriers (69%), compared with S4, S1, and S2 (56%, 44%, and 47%). Medical history profiles also showed the highest prevalence of cardiovascular conditions in S1 (21.8%) and the highest prevalence of smoking history in S4 (47.9%), consistent with the subtype rankings in ADNI (**Figure 6D**). Additional demographic characteristics among the four subtypes in the NACC cohort are in **Supplementary Table S3**.

**Figure 6.**
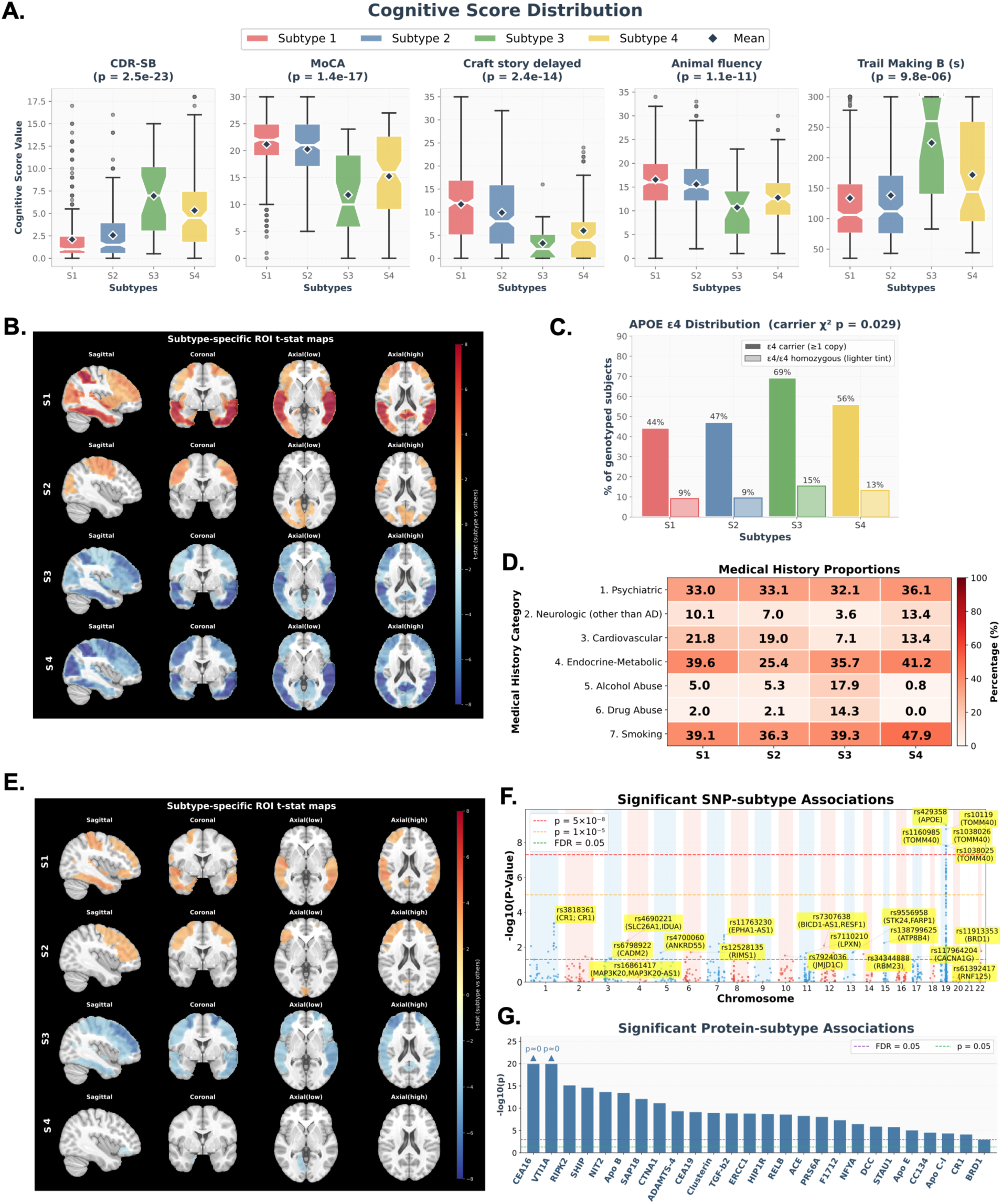
External validation of ADSMoE-derived multimodal AD subtypes in the NACC and UKB cohorts. (**A-D**) Subtype characterization in the NACC cohort. (**A**) Distributions of five cognitive assessments across four subtypes (S1-S4), including CDR-SB, MoCA, delayed story recall, animal fluency, and Trail Making Test B. Diamonds indicate mean values, and P values represent overall differences across subtypes. (**B**) Subtype-specific regional brain volume patterns visualized using ROI-level Welch’s t-statistic, comparing each subtype with the remaining subtypes. (**C**) Distribution of APOE ε4 carriers and homozygous individuals across subtypes. (**D**) Prevalence of medical history characteristics across subtypes. (**E-G**) Subtype characterization in the UKB cohort. (**E**) Subtype-specific regional brain volume patterns visualized using ROI-level Welch’s t-statistic. (**F**) Manhattan plot of SNP-subtype associations across chromosomes, highlighting significant and suggestive genetic loci. (**G**) Subtype-associated proteins ranked by association significance (−log₁₀(P)), with nominal and false discovery rate significance thresholds indicated.

#### Subtype characterization in the UKB cohort

We further applied our proposed and trained ADSMoE to the independent UKB cohort without retraining or fine-tuning to evaluate the cross-cohort generalizability of the identified AD subtypes. Neuroimaging analyses revealed neuroanatomical patterns consistent with those observed in ADNI and NACC (**Figure 6E**), with S1 showing predominantly positive ROI t-statistics and S3 exhibiting widespread negative values across brain regions. Genetic association analyses identified prominent signals on chromosome 19 (**Figure 6F**), particularly within the *APOE* and *TOMM40* regions, aligned with the genetic associations observed in ADNI cohort. Additional associations were also identified at established AD susceptibility loci, including *rs3818361* in *CR1* and *rs11763230* in the *EPHA1-AS1* region. Proteomic association analyses also uncovered significant molecular differences among subtypes (**Figure 6G**), with *CEA16* and *VTI1A* showing the strongest associations, along with several proteins also identified in ADNI, such as *STAU1, DCC, VTI1A,* and *Apo B*. These findings demonstrate that ADSMoE-derived subtypes retain consistent neuroanatomical, genetic, and proteomic characteristics across independent cohorts, supporting their biological relevance and the generalizability of the framework to heterogeneous multimodal datasets. Additional demographic characteristics among the four subtypes in UKB cohort are provided in **Supplementary Table S4**.

## Discussion

In this study, we developed ADSMoE, a flexible multimodal framework for discovering AD subtypes from heterogeneous real-world data via expert-based representation learning. A major challenge in real-world AD research is that multimodal data are inherently heterogeneous and frequently incomplete, resulting in the exclusion of large numbers of participants when conventional multimodal methods require complete observations or a single representation pathway. ADSMoE addresses this challenge by leveraging all available modalities without extensive imputation or discarding subjects. By combining flexible multimodal encoding with a subtype-specific MoE decoder, the framework accommodates partially observed multimodal data while capturing distinct disease patterns. Unlike conventional subtype discovery methods that rely on predefined disease categories or modality-specific clustering strategies, ADSMoE learns disease subtypes as emergent properties of multimodal representation learning. More importantly, expert specialization enables the model to preserve heterogeneous disease patterns rather than collapsing disease heterogeneity into a shared representation.

ADSMoE demonstrates robust, reproducible multimodal representation learning and subtype discovery across model configurations and experimental conditions. The Fleximodal MoE-based encoder achieved a favorable balance between predictive performance and computational efficiency. Furthermore, ADSMoE identified stronger subtype-genetic associations while maintaining competitive clustering quality. Sensitivity analyses further showed that the learned subtype structure remained stable across a broad range of model configurations, indicating that subtype discovery was not driven by specific hyperparameter choices. This robustness likely arises from the complementary effects of normative modeling, deep representation learning, and expert specialization, which together enhance latent representation stability and separability. Additionally, compared with Gene-SGAN’s genetically associated imaging subtyping, ADSMoE advances toward a unified multimodal framework for AD subtyping. These findings indicate that ADSMoE learns robust and biologically relevant multimodal representations, providing a foundation for modeling disease heterogeneity in AD.

A notable finding of this study is the convergence of multiple data modalities toward a coherent four-subtype organization. The identified subtypes exhibited consistent differences across structural neuroimaging, genetic variation, protein biomarkers, clinical characteristics, and survival rates from CN to AD, supporting their biological validity. Subtype 1 was characterized by relatively preserved baseline cognition, higher cortical volumes, and lower p-tau levels than other subtypes, resembling a minimal-atrophy phenotype, yet showed faster long-term progression to AD. Subtype 2 represented an older subgroup with moderate cognitive impairment, relatively preserved neocortical volumes, showing potential similarities to a limbic-predominant AD phenotype^67^. Subtype 3 exhibited the highest PRS, the most severe cognitive impairment, and the most extensive cortical atrophy, accompanied by elevated expression of lipid-associated proteins, indicating a typical AD neuroanatomical pattern^67^. Lastly, Subtype 4 showed intermediate cognitive impairment with relatively higher temporal cortical volumes and lower volumes in portions of the frontal, parietal, and occipital cortices. It was also distinguished by limited genetic risk signals and lower expression of AD- and inflammation-related proteins, including *Apo C-I* and *CEA16*. Together with its comparatively slower clinical progression, these findings support a resilience-associated subtype in which preserved brain integrity and reduced molecular burden may mitigate disease progression^68^.

The biological relevance of the identified subtypes is further supported by their associations with established AD-related genetic variants and protein biomarkers. Among all SNPs associated with the identified subtypes, variants within the *APOE* locus, including *rs429358* and *rs7412*, showed the strongest subtype-specific associations, consistent with their well-established roles in AD susceptibility, brain atrophy and cognitive decline^69^. Additional subtype–associated variants mapped to genes involved in amyloid processing (*CLSTN1*)^70^, tau secretion (*FRMD4A*)^71^, microglial activation (*MS4A3*)^72^, endocytosis (*BIN1*)^73^, and neuroinflammatory pathways (*SEMA3F*, *MEF2C*, *HLA-DQA1*, and *LAT*)^74^, indicating that distinct biological mechanisms contribute to different subtype profiles. Similarly, subtype-enriched proteins, such as *STAU1*, *DCC*, *VTI1A*, and *ApoB*^75,76^, participate in protein transport, immune regulation, and lipid metabolism, further supporting subtype–specific molecular signatures. Collectively, these genetic and proteomic associations indicate that the identified subtypes are underpinned by distinct biological pathways, supporting the ability of ADSMoE to uncover mechanistically meaningful disease heterogeneity.

Importantly, the generalizability of ADSMoE was further supported by external validation using independent NACC and UKB cohorts. Despite differences in multimodal data availability, the identified subtypes retained several clinical and biological characteristics observed in ADNI. In NACC, Subtype 3 consistently exhibited greater cognitive impairment, widespread reductions in regional brain volumes, and the highest prevalence of APOE ε4 carriage, indicating the consistency of its clinical, neuroanatomical and genetic characteristics across cohorts. Similarly, MRI analyses in UKB revealed comparable subtype-specific patterns. Genetic analyses further identified prominent associations within the *APOE* region, consistent with those observed in ADNI, alongside additional signals involving established AD susceptibility loci (e.g., *CR1*). Proteomic analyses further revealed overlapping subtype-associated proteins, including *STAU1*, *VTI1A*, *DCC*, *CEA16*, and *ApoB*, although molecular profiles differed between cohorts. These findings suggest that ADSMoE can transfer learned multimodal representations across independent cohorts, while preserving clinically and biologically meaningful subtype characteristics, supporting its potential for generalizable AD heterogeneity modeling.

This study has several limitations. First, although the framework was evaluated across three independent cohorts, further validation in larger and more diverse populations and healthcare systems are needed to establish its robustness and generalizability. Second, our cross-cohort harmonization did not fully account for platform- and biospecimen-specific differences, particularly between CSF and plasma proteomic measurements, potentially affecting the comparability of molecular associations. Third, while the identified subtypes demonstrated distinct longitudinal clinical trajectories, subtype discovery primarily relied on cross-sectional multimodal measurements, and incorporating longitudinal multimodal data may further improve subtype resolution and capture dynamic changes in disease progression. Fourth, although the identified genetic and proteomic associations support the biological relevance of the discovered subtypes, experimental validation is needed to establish their mechanisms and potential causal contributions to AD progression. Finally, integrating additional data modalities, such as plasma multi-omics, digital biomarkers, and wearable sensor data, may further enhance subtype characterization and support future precision medicine applications.

In summary, this study presents a unified, multimodal representation learning framework for characterizing AD heterogeneity using real–world data. By jointly learning disease subtypes as multimodal signatures through a MoE architecture, ADSMoE accommodates incomplete modality combinations and enables biologically informed patient stratification. Across three independent AD cohorts, the four identified subtypes exhibited consistent differences in neuroanatomical characteristics, genetic and proteomic associations, and clinical profiles, providing evidence for their biological and clinical relevance. More broadly, the proposed framework provides a scalable foundation for subtype-specific biomarker discovery, precision clinical trial design, and mechanism-guided therapeutic development for AD and other neurodegenerative disorders.

## Methods

### Data sources and study cohorts

We collected multimodal data, including neuroimaging, genetic, clinical, and proteomic measurements, from three large-scale cohorts: the Alzheimer’s Disease Neuroimaging Initiative (ADNI), the National Alzheimer’s Coordinating Center (NACC), and the UK Biobank (UKB). ADNI data were downloaded on June 15, 2025. The study included 5,422 participants across the three cohorts (**Supplementary Table S1**). Participants with AD and CN were identified using ADNI diagnostic records (DXSUM)^77^, NACC clinical assessments^78^, and UKB health records (ICD-10 codes for AD)^79^. For external validation, we randomly selected eligible participants with AD from NACC and UKB to obtain sample sizes equal to that of the ADNI cohort. Random sampling was performed independently within each external cohort using a fixed random seed of 42. For each participant, we selected the image, clinical, and proteomic data to improve temporal alignment across modalities. CN participants served as reference populations for normative modeling to account for disease-unrelated variations and identify deviations from healthy reference. The ADNI cohort was used for model development and training, whereas NACC and UKB were reserved for independent external validation cohorts without model retraining or fine-tuning.

### Multimodal data processing

#### Image data processing

T1-weighted structural MRI scans were obtained from ADNI and from a previously curated NACC MRI dataset described by Pan et al.^80^ These scans were then processed using a fully automated and standardized FreeSurfer (version 7.2.0) pipeline. The processing pipeline included N3 bias field correction, followed by anatomical registration and intensity normalization. Then, extracranial tissues were removed via automated skull stripping. The skull-stripped images then underwent white matter segmentation and cortical surface reconstruction. Anatomical parcellation was performed using the Desikan-Killiany (DK) atlas^81^, while subcortical structures were automatically segmented. The resulting regional morphometric features were then extracted from the FreeSurfer outputs. For the UKB cohort, we used precomputed imaging-derived phenotypes (IDPs) generated using FreeSurfer, including cortical measures based on the DK white-surface parcellation and volumetric measures derived from automated segmentation. All imaging features were stored in a subject-level matrix indexed by participant identifiers.

#### Genetics data processing

Genetic features were derived from whole-genome sequencing (WGS) data in ADNI and imputed genotype data in the UKB. To improve cross-cohort consistency and reduce the feature space, analyses in both datasets were restricted to a curated set of AD-associated SNPs obtained from the GWAS Catalog (N = 5,343)^82^. For ADNI, per-chromosome variant call format (VCF) files from WGS data were processed, and genome-wide variant quality control was performed on autosomal chromosomes (1–22) using vcftools^83^, including filters on minor allele frequency (MAF ≥ 0.01), per-variant call rate (≥ 0.95), and Hardy-Weinberg equilibrium (HWE P ≥ 1×10⁻⁶). Variants with insertion or deletion alleles were excluded. Following quality control, variants were further restricted to the curated AD-associated SNP list. For the UKB, imputed genotype data from the imputation release were used in BGEN format. The same curated SNP list was applied by extracting variants from chromosome-wide BGEN files using bgenix, followed by conversion to PLINK2^84^ format and restriction to participants to the analytic cohort. Imputed genotypes were hard-called using a fixed probability threshold of 0.1 and exported as compressed VCF files. For both cohorts, filtered VCFs were processed with scikit-allel^85^, and diploid genotypes were converted into an additive allele-count representation (0/1/2) based on the number of alternative alleles, yielding subject-by-variant matrices indexed by participant identifiers, which served as genetic feature inputs.

#### Clinical data processing

Clinical features were derived from routinely collected phenotypic data in each cohort, including demographic characteristics, neurological examinations, medical history, and vital signs. Prior to modeling, non-informative variants (e.g., participant identifiers) and common study-specific missing codes (e.g., −4) were removed or treated as missing. For measurements reported in different units, values were converted to a consistent unit system before feature construction. Continuous variables were then normalized using a standard scaler to ensure comparable numerical scales, whereas categorical variables were transformed into binary indicators using one-hot encoding. To avoid unstable high-dimensional expansions, categorical fields with high cardinality (>=20) were excluded. After preprocessing, continuous and one–hot encoded categorical variables were combined into participant–level clinical feature matrix as model input.

#### Proteomic data processing

Protein abundance measurements were used as the proteomic modality. For ADNI, CSF protein measurements generated using the SomaScan v4.1 aptamer-based proteomics platform (∼7,000 protein targets) were used in the form of relative fluorescence unit (RFU) values. For UKB, plasma protein measurements generated using the Olink Explore proteomics platform (∼3,000 protein targets) were analyzed using the provided normalized protein expression (NPX) values. Within each cohort, proteins with more than 20% missing values or near-zero variance across participants were excluded. The remaining protein measurements were log-transformed where appropriate and standardized using z-score normalization to ensure comparable numerical scales.

#### Cross-cohort harmonization

To improve comparability across ADNI, NACC, and UKB, modality-specific harmonization procedures were implemented using shared features identified across cohorts. Features with more than 20% missing values or low variance (variance < 0.1) were excluded. Missing values in the remaining continuous features were imputed using a k-nearest neighbors (KNN) imputer (k = 5). Missingness in categorical variables was encoded using a unified missing code. Modality-specific harmonization procedures were applied as follows. For image data, 189 shared MRI features were identified across cohorts, and ComBat harmonization^86^ was applied using ADNI as the reference batch, yielding a harmonized imaging matrix across the three cohorts. For clinical data, continuous variables (e.g., vital signs) were harmonized using ComBat and subsequently combined with the remaining encoded categorical variables, resulting in a merged clinical feature matrix containing 57 variables. For genetic data, variants were merged across cohorts based on shared reference SNP identifiers (rsIDs), yielding a harmonized genetic feature matrix comprising 937 SNPs. For proteomic data, protein measurements were standardized to z-scores using StandardScaler, followed by ComBat harmonization to account for cohort-specific differences. A total of 126 matched proteins were used for downstream modeling.

### Normative modeling for disease-related feature extraction

To reduce the effects of demographic and other non-disease-related variations, we applied normative modeling^49,50^ using CN participants as the reference population. For each feature *j*, a linear regression model

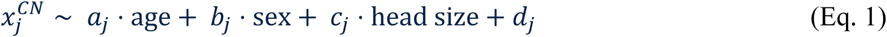

was fitted on CN data using age, sex, and head size, where *a*_j_, *b*_j_, and *c*_j_ are the regression coefficients for age, sex, and head size, respectively, and *d*_j_ is the intercept. Age, sex, and head size are commonly included as covariates in neuroimaging studies^87,88^, with head size typically measured by intracranial volume (ICV) to account for interindividual differences in regional brain volumes. The fitted coefficients were then used to estimate the expected feature value for each AD patient. For each feature *j* of AD patient *i* , the standardized residual *r_i_*_j_ was calculated as the difference between the observed *x_i_*_j_ and expected *x̂_j_* (*age_i_*, *sex_i_*, ℎ*ead_i_*) divided by the standard deviation in CN, *σ*_j_(*CN*):

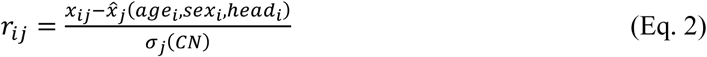

The resulting residuals from image, proteomic, and clinical features, together with original genetic SNP features without residualization, were used as multimodal inputs to ADSMoE. Additional details of the normative modeling procedure are provided in **Supplementary Figure S5**.

### Multimodal representation learning using Fleximodal encoder

Heterogeneous and incomplete multimodal input poses a practical challenge for learning a unified latent space, as conventional early-fusion approaches typically require complete inputs^89^, while zero-padding or extensive imputation may introduce bias. To address this, we adopted an autoencoder architecture with a Fleximodal encoder^47^, which accommodates arbitrary combinations of available modalities and is adapted from recent Mixture-of-Experts (MoE)-based multimodal learning frameworks^47^. Specifically, each observed modality *m* was first mapped to a shared embedding space *e_i_^m^* by a modality-specific encoder for participant *i*. For missing modalities, the corresponding embeddings were replaced with learnable tokens from a missing-modality bank: *e_i_^m^* = Encoder_m_(*i*) if modality *m* is observed, otherwise *B_m_* . The completed multimodal token *x* was then fused by an MoE layer (used as the feed-forward block within a lightweight Ttransformer), where a router selects the top-*K* experts and outputs a weighted mixture: *y* = 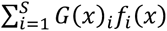, with *G*(*x*) = TopK NsoftmaxO*g*(*x*)PQ and *g*(⋅) implemented as an MLP classifier. We adapted a two-stage routing strategy for training: a generalized router was first trained using fully observed samples to initialize expert representations, and a specialized router was then trained using samples with missing modalities to promote expert specialization across different modality combinations. During the multimodal representation learning, the Fleximodal encoder and decoder were jointly optimized, with the decoder employing default top-*K* expert routing. Following training, the encoded tokens of each patient were aggregated using masked mean pooling over observed modalities to obtain a latent vector. The resulting latent representations were used for subtype initialization and subsequent joint optimization in ADSMoE.

### Subtype initialization via expectation-maximization (EM)

To provide a stable initialization for subtype-specific expert routing, we applied an EM algorithm^48^ to identify initial AD subtype groups. EM is an iterative method that estimates cluster assignments and model parameters by alternating between expectation and maximization steps. Using the pretrained Fleximodal encoder, we obtained a latent representation ℎ*_i_* for each participant with AD. For each EM cluster *k*, the mean latent representation *μ_k_* was calculated and used to initialize the corresponding decoder gate weight vector as:

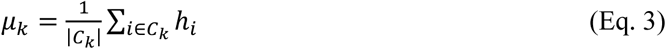

where *C_k_* denotes the set of AD participants assigned to EM cluster *k*. The decoder gate then computed patient-level soft routing probabilities based on the cosine similarity between the centered latent representation and the initialized gate weights:

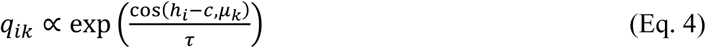

where *q_ik_* denotes the routing probability of patient *i* to subtype expert *k* , and *τ* is the temperature parameter controlling the sharpness of the routing distribution. The resulting probabilities were used to initialize subtype-specific expert routing for subsequent joint optimization.

### Multimodal AD subtype discovery using the MoE-based decoder

#### Expert specialization

After encoder pretraining and subtype initialization, multimodal AD subtyping was learned through a subtype-specific MoE decoder that reconstructs multimodal inputs while allowing different experts to capture subtype-specific characteristics. Specifically, the decoder takes modality-wise latent tokens from the Fleximodal encoder. As shown in **Supplementary Figure S5,** the feed-forward network (FFN) in the first transformer layer is replaced by an MoE module containing *K* subtype-specific experts. Unlike the default top-*K* routing used during pretraining, the decoder employs patient-level soft routing when learning subtypes, where all experts contribute to the reconstruction. Specifically, the decoder combines expert outputs using the routing probabilities as 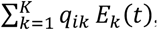, where *E_k_*(*t*) denotes the output of subtype expert *k* for token *t*. This mechanism allows different experts to specialize in distinct multimodal disease patterns. Additionally, to account for disease severity and reduce its influence on subtype-specific representations, we introduced a separate severity head that generates a positive severity score *s_i_* for each patient from the pooled latent representation, computed as softplus(*w* ⋅ ℎ*_i_* + *b*), where *w* and *b* are learnable parameters. The expert outputs are then processed by a transformer layer and scaled by the severity score *s_i_*, followed by modality-specific projections to reconstruct the original input features. Reconstruction loss was calculated using mean squared error (MSE) denoted as:

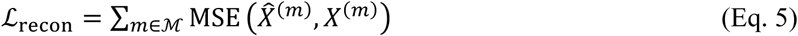

where *x̂_(m)_* and *X*^(*m*)^ denote the reconstructed and target features of modality (*m*), and ℳ denotes the set of modalities included in reconstruction loss. During the representation learning phase, learnable embeddings were used as reconstruction targets for missing modalities, whereas during the subtype discovery phase, reconstruction loss was calculated only for observed modalities. To reduce the risk of routing collapse, we applied an expert-balancing regularization term that encourages approximately uniform expert utilization across patients. The overall joint optimization objective combines reconstruction loss and expert-balancing regularization as

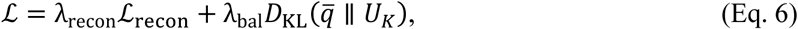

where *q̄* is the mean routing probability distribution across patients, *D*_KL_ is the Kullback–Leibler divergence, *U_K_* denotes the uniform distribution over K subtype experts, and the weighting coefficients were λ_recon_and λ_bal_. During this phase, the encoder, decoder, and routing network were jointly optimized to learn subtype-specific multimodal representations.

#### Subtype assignment

Final subtype discovery was determined after model training. For each participant with AD, the trained decoder generated routing probabilities *q_k_* across the *K* subtype-specific experts, and the subtype with the highest routing probability was assigned as the initial label. We set *K* = 4, consistent with previous studies identifying four distinct neuroimaging-based AD subtypes^90^. To assess the stability of identified subtypes, model training was repeated multiple times with different random seeds comprising at least 80% of participants. Because expert indices are arbitrary across training runs, subtype labels were aligned to a reference run using Hungarian matching algorithm^91^. The final subtype of each AD participant was determined by majority voting across aligned runs, and the assignment confidence was quantified as the proportion of runs agreeing with the final subtype label.

### Model evaluation and experimental settings

#### Model comparison

To evaluate multimodal representation learning, we compared three MoE-based encoders: FlexMoE (Flexible Mixture-of-Experts)^47^, LiMoE (Language-Image Mixture of Experts)^42^, and FuseMoE (Mixture-of-Experts Transformers for Fleximodal Fusion)^41^, and a conventional convolutional neural network ResNet^92^. FlexMoE and FuseMoE are specifically designed for flexible multimodal learning. We used AD vs. CN classification as a benchmark task to assess the discriminative capability of the learned disease-relevant multimodal representations. For AD subtype discovery, we compared ADSMoE with representative classical and deep clustering methods, including GeneSGAN (Gene-guided weakly-supervised clustering via generative adversarial networks)^29^, CDEC (contrastive deep embedded clustering)^93^, EDESC (efficient deep embedded subspace clustering)^94^, K-means^95^ and spectral clustering^96^. These approaches represent different strategies for integrating representation learning with clustering or identifying latent patient subgroups. Model comparisons were conducted under multiple experimental settings to assess robustness to feature dimensionality and incomplete multimodal observations. Specifically, we evaluated (1) varying proportions of genetic features (i.e., 50%, 75%, and 100% of the SNP feature set); (2) missingness in selected neuroimaging regions (i.e., missingness of entorhinal, hippocampus, precuneus features); (3) missingness across different categories of clinical data (e.g., demographics, medical history and neurological assessments); and (4) varying proportions of proteomic features (i.e., 50%, 75% and 100% of the protein feature set). These experiments were designed to examine the sensitivity of ADSMoE and competing methods to changes in feature availability across modalities.

#### Evaluation metrics

We evaluated models in two settings. First, to assess multimodal representation learning, we reported binary classification metrics on the validation and test sets, including accuracy (Acc), F1–score (F1), and the area under the receiver operating characteristic curve (AUC). Computational efficiency was evaluated using wall-clock runtime (seconds). Second, to evaluate AD subtyping performance (**Figure 2B**), we assessed clustering quality using the Davies-Bouldin index (DBI)^54^, with lower values indicating more compact and better-separated clusters. Additionally, to evaluate the biological relevance of the identified subtypes, we quantified genetic associations using the number of subtype-associated single-nucleotide polymorphisms (N-Asso-SNPs) with AD. Following prior imaging–genetics subtyping studies (i.e., GeneSGAN), SNP-subtype associations were assessed using likelihood-ratio tests (LRTs) comparing multinomial models with and without each SNP. N–Asso–SNPs was defined as the number of SNPs with LRT p < 0.05 (K − 1 degrees of freedom).

#### Experimental settings

The model was implemented in PyTorch and trained on a single NVIDIA L4 GPU (24 GB), with the three-phase training pipeline taking approximately 3 minutes per run. Performance was assessed using five-fold cross-validation. The encoder and decoder used 16 tokens per modality, a hidden dimension of 128, and 8 attention heads, with 3 and 2 transformer blocks, respectively. The encoder employed 16 experts with top-2 routing, while the decoder used 4 subtype experts with soft mixture routing (dropout = 0.1). We optimized the model using Adam (learning rate 1×10^-4^, weight decay 1×10^-5^, batch size 32): 40 epochs of multimodal representation learning, followed by 30 epochs of joint training. After training on ADNI, the model was frozen and applied to NACC and UKB using cohort-specific residuals, without retraining or fine-tuning. More hyperparameter settings are provided in **Supplementary Table S5**.

### Statistical Analysis

To characterize subtype-specific neuroanatomical patterns, ROI-level differences were assessed using two-sample Welch’s t-tests^97^, comparing each subtype with all remaining participants. ROIs were ranked by the absolute t-statistic (|*t*|) for visualization. To test SNP-subtype associations, LRTs were performed using multinomial logistic regression models with subtype labels as the dependent variable and SNP genotype as the predictor. Models were fitted using the limited-memory Broyden-Fletcher-Goldfarb-Shanno (L-BFGS)^98^ optimizer with a maximum of 1,000 iterations. The LRT statistic was evaluated using a chi-square distribution with *K*-1 degrees of freedom, where *K* denotes the number of subtypes. Genetic association analyses were adjusted for age and sex. Additionally, subtype-specific proteomic associations were assessed by comparing protein measurements across the identified subtypes using one-way analysis of variance (ANOVA), with an omnibus F-test applied across all subtypes. Multiple testing was controlled using the Benjamini-Hochberg false discovery rate^99^ (FDR; q < 0.05). For genetic associations, the conventional genome-wide significance threshold (*P* = 5 × 10⁻⁸) and suggestive significance threshold (*P* = 1 × 10⁻⁵) were also used as reference threshold. For longitudinal analyses, time to AD diagnosis was defined as the interval in years between the baseline assessment and the first recorded AD diagnosis. Subtype labels assigned using data from the first recorded AD diagnosis were linked retrospectively to each participant’s earlier CN or MCI visit. Only participants with an observed CN-to-AD or MCI-to-AD transition were included, so there was no right censoring. Kaplan-Meier survival curves^100^ were estimated using the lifelines package, and subtype differences were assessed using a multivariate log-rank test. Conversion times exceeding 20 years were excluded from the analysis. Cross-modal associations were visualized using a DIABLO-based Circos plot, retaining component-based similarities with absolute values greater than 0.4.

## Data availability

The data for this study include ADNI (https://adni.loni.usc.edu/), NACC (https://www.naccdata.org/), and UKB (https://www.ukbiobank.ac.uk/). Application processes for these data sources include acceptance of the Data Use Agreement and submission of an online application form. Detailed instructions for data access and request procedures can be found on the corresponding websites. AD-relevant SNPs were pre-selected using GWAS summary from GWAS-Catalog (https://www.ebi.ac.uk/gwas/efotraits/MONDO_0004975).

## Code availability

All codes are available at https://github.com/UF-HOBI-Yin-Lab/Multimodal_AD_Subtyping_ADSMoE.

## Acknowledgements

This study is supported by the Florida Department of Health Ed and Ethel Moore Alzheimer’s Disease Research Program (26A11). Data used in this study were obtained in part from the Alzheimer’s Disease Neuroimaging Initiative (ADNI). Data collection and sharing for the Alzheimer’s Disease Neuroimaging Initiative (ADNI) is funded by the National Institute on Aging (National Institutes of Health Grant U19AG024904). The grantee organization is the Northern California Institute for Research and Education. In the past, ADNI has also received funding from the National Institute of Biomedical Imaging and Bioengineering, the Canadian Institutes of Health Research, and private sector contributions through the Foundation for the National Institutes of Health (FNIH) including generous contributions from the following: AbbVie, Alzheimer’s Association; Alzheimer’s Drug Discovery Foundation; Araclon Biotech; BioClinica, Inc.; Biogen; BristolMyers Squibb Company; CereSpir, Inc.; Cogstate; Eisai Inc.; Elan Pharmaceuticals, Inc.; Eli Lilly and Company; EuroImmun; F. Hoffmann-La Roche Ltd and its affiliated company Genentech, Inc.; Fujirebio; GE Healthcare; IXICO Ltd.; Janssen Alzheimer Immunotherapy Research & Development, LLC.; Johnson & Johnson Pharmaceutical Research & Development LLC.; Lumosity; Lundbeck; Merck & Co., Inc.; Meso Scale Diagnostics, LLC.; NeuroRx Research; Neurotrack Technologies; Novartis Pharmaceuticals Corporation; Pfizer Inc.; Piramal Imaging; Servier; Takeda Pharmaceutical Company; and Transition Therapeutics. The NACC database is funded by NIA/NIH Grant U24 AG072122. NACC data are contributed by the NIA-funded ADRCs: P30 AG062429 (PI James Brewer, MD, PhD), P30 AG066468 (PI Oscar Lopez, MD), P30 AG062421 (PI Teresa Gomez-Isla, MD), P30 AG066509 (PI Thomas Grabowski, MD), P30 AG066514 (PI Mary Sano, PhD), P30 AG066530 (PI Helena Chui, MD, Arthur Toga, PhD), P30 AG066507 (PI Marilyn Albert, PhD), P30 AG066444 (PI David Holtzman, MD), P30 AG066518 (PIs Lisa Silbert, MD, Kevin Duff, PhD), P30 AG066512 (PI Thomas Wisniewski, MD), P30 AG066462 (PI Scott Small, MD), P30 AG072979 (PI David Wolk, MD), P30 AG072972 (PIs Charles DeCarli, MD, Rachel Whitmer, PhD), P30 AG072976 (PI Andrew Saykin, PsyD), P30 AG072975 (PI Julie Schneider, MD, MS), P30 AG072978 (PI Ann McKee, MD), P30 AG072977 (PI Robert Vassar, PhD), P30 AG066519 (PI Joshua Grill, PhD), P30 AG062677 (PIs Brad Boeve, MD, Ronald Petersen, MD, PhD), P30 AG079280 (PI Jessica Langbaum, PhD), P30 AG062422 (PI Gil Rabinovici, MD), P30 AG066511 (PI Allan Levey, MD, PhD), P30 AG072946 (PI Linda Van Eldik, PhD), P30 AG062715 (PI Sanjay Asthana, MD, FRCP), P30 AG072973 (PI Russell Swerdlow, MD), P30 AG066506 (PIs Glenn Smith, PhD, ABPP, David Lowenstein, PhD, Ranjan Duara, MD), P30 AG066508 (PIs Stephen Strittmatter, MD, PhD, Christopher Van Dyck, MD), P30 AG066515 (PI Victor Henderson, MD, MS), P30 AG072947 (PI Suzanne Craft, PhD), P30 AG072931 (PI Henry Paulson, MD, PhD), P30 AG066546 (PIs Sudha Seshadri, MD, Gladys Maestre, MD, PhD), P30 AG086401 (PI Erik Roberson, MD, PhD), P30 AG086404 (PI Gary Rosenberg, MD), P30 AG086403 (PI Angela Jefferson, PhD), P30 AG072958 (PIs Heather Whitson, MD, Gwenn Garden, MD, PhD), P30 AG072959 (PI Jagan Pillai, MD, PhD), P30 AG092752 (Ihab Hajjar, MD, MS). This research has been conducted using the UK Biobank Resource under Application Number 95030.

## Contributions

W.M. and R.Y. conceived ideas and developed the initial concept. W.M. designed the experiments, analytic plan, and codes. W.M. and J.P. conducted data collection, processing, and result interpretation. W.M. and M.Y. performed the experiments. W.M. wrote the manuscript with support from R.Y., J.P., M.Y., P.Z., K.G., M.L., J.X., M.B., Y.W., and R.Y. provided critical feedback and helped shape the research, analysis, and manuscript. R.Y. supervised the project.

## References

1. 2025 Alzheimer’s disease facts and figures. Alzheimers. Dement. 21, (2025).

2. Dubois, B. et al. Preclinical Alzheimer’s disease: Definition, natural history, and diagnostic criteria. Alzheimers. Dement. 12, 292–323 (2016).

3. Hunter, T. R., Santos, L. E., Tovar-Moll, F. & De Felice, F. G. Alzheimer’s disease biomarkers and their current use in clinical research and practice. Mol. Psychiatry 30, 272–284 (2025).

4. Malarte, M.-L. et al. Discriminative binding of tau PET tracers PI2620, MK6240 and RO948 in Alzheimer’s disease, corticobasal degeneration and progressive supranuclear palsy brains. Mol. Psychiatry 28, 1272–1283 (2023).

5. Jack, C. R., Jr et al. NIA-AA Research Framework: Toward a biological definition of Alzheimer’s disease. Alzheimers. Dement. 14, 535–562 (2018).

6. Dickerson, B. C. et al. Alzheimer-signature MRI biomarker predicts AD dementia in cognitively normal adults. Neurology 76, 1395–1402 (2011).

7. Hansson, O., Blennow, K., Zetterberg, H. & Dage, J. Blood biomarkers for Alzheimer’s disease in clinical practice and trials. *Nat*. Aging 3, 506–519 (2023).

8. Salvadó, G. et al. Disease staging of Alzheimer’s disease using a CSF-based biomarker model. *Nat*. Aging 4, 694–708 (2024).

9. Shi, Y. & Holtzman, D. M. Interplay between innate immunity and Alzheimer disease: APOE and TREM2 in the spotlight. Nat. Rev. Immunol. 18, 759–772 (2018).

10. Bertram, L. & Tanzi, R. E. Thirty years of Alzheimer’s disease genetics: the implications of systematic meta-analyses. Nat. Rev. Neurosci. 9, 768–778 (2008).

11. Bird, T. D. Alzheimer Disease Overview. (University of Washington, Seattle, Seattle, 1993).

12. Elliott, L. T. et al. Genome-wide association studies of brain imaging phenotypes in UK Biobank. Nature 562, 210–216 (2018).

13. Wen, J. et al. Genomic loci influence patterns of structural covariance in the human brain. Proc. Natl. Acad. Sci. U. S. A. 120, e2300842120 (2023).

14. Bucci, M., Chiotis, K., Nordberg, A. & Alzheimer’s Disease Neuroimaging Initiative. Alzheimer’s disease profiled by fluid and imaging markers: tau PET best predicts cognitive decline. Mol. Psychiatry 26, 5888–5898 (2021).

15. Stevenson-Hoare, J. et al. Plasma biomarkers and genetics in the diagnosis and prediction of Alzheimer’s disease. Brain 146, 690–699 (2023).

16. Kim, S. et al. Genome-wide association study of CSF biomarkers Abeta1-42, t-tau, and p-tau181p in the ADNI cohort. Neurology 76, 69–79 (2011).

17. Acosta, J. N., Falcone, G. J., Rajpurkar, P. & Topol, E. J. Multimodal biomedical AI. Nat. Med. 28, 1773–1784 (2022).

18. Zhang, Y., et al. MmFormer: Multimodal medical transformer for incomplete multimodal learning of brain tumor segmentation. in Lecture Notes in Computer Science 107–117 (Springer Nature Switzerland, Cham, 2022).

19. Wang, H. et al. Multi-modal learning with missing modality via shared-specific feature modelling. in 2023 IEEE/CVF Conference on Computer Vision and Pattern Recognition (CVPR) (IEEE, 2023). doi:10.1109/cvpr52729.2023.01524.

20. Estarellas, M., Oxtoby, N. P., Schott, J. M., Alexander, D. C. & Young, A. L. Multimodal subtypes identified in Alzheimer’s Disease Neuroimaging Initiative participants by missing-data-enabled subtype and stage inference. Brain Commun. 6, fcae219 (2024).

21. Veitch, D. P. et al. The Alzheimer’s Disease Neuroimaging Initiative in the era of Alzheimer’s disease treatment: A review of ADNI studies from 2021 to 2022. Alzheimers. Dement. 20, 652–694 (2024).

22. Aisen, P. S. et al. Clinical Core of the Alzheimer’s Disease Neuroimaging Initiative: progress and plans. Alzheimers. Dement. 6, 239–246 (2010).

23. Huang, Y. et al. Identifying Alzheimer’s disease progression subphenotypes via a graph-based framework using electronic health records. *J*. Healthc. Inform. Res. 10, 317–339 (2026).

24. Meng, W. et al. An interpretable population graph network to identify rapid progression of Alzheimer’s disease using UK Biobank. in (AMIA Annual Symposium Proceeding Archive, 2024).

25. Xu, J. et al. Identification of Outcome-Oriented Progression Subtypes from Mild Cognitive Impairment to Alzheimer’s Disease Using Electronic Health Records. in (AMIA Annual Symposium Proceedings Archive, 2023).

26. Young, A. L. et al. Uncovering the heterogeneity and temporal complexity of neurodegenerative diseases with Subtype and Stage Inference. Nat. Commun. 9, 4273 (2018).

27. Young, A. L. et al. Ordinal SuStaIn: Subtype and Stage Inference for clinical scores, visual ratings, and other ordinal data. *Front*. Artif. Intell. 4, 613261 (2021).

28. Tandon, R., Lah, J. J. & Mitchell, C. S. s-SuStaIn: Scaling subtype and stage inference via simultaneous clustering of subjects and biomarkers. Proc. Mach. Learn. Res. 248, 461–476 (2024).

29. Yang, Z. et al. Gene-SGAN: discovering disease subtypes with imaging and genetic signatures via multi-view weakly-supervised deep clustering. Nat. Commun. 15, 354 (2024).

30. Jiao, C.-N., Gao, Y.-L., Ge, D.-H., Shang, J. & Liu, J.-X. Multi-modal imaging genetics data fusion by deep auto-encoder and self-representation network for Alzheimer’s disease diagnosis and biomarkers extraction. Eng. Appl. Artif. Intell. 130, 107782 (2024).

31. Afxenti, S. et al. Integrating imaging and omics for enhanced subtyping of mild cognitive impairment associated with Alzheimer’s disease. J. Transl. Med. 23, 1240 (2025).

32. Machado Reyes, D., et al. Identifying progression-specific Alzheimer’s subtypes using multimodal transformer. J. Pers. Med. 14, 421 (2024).

33. Xu, E. et al. Pathology steered stratification network for subtype identification in Alzheimer’s disease. Med. Phys. 51, 1190–1202 (2024).

34. Varol, E., Sotiras, A. & Davatzikos, C. HYDRA: Revealing heterogeneity of imaging and genetic patterns through a multiple max-margin discriminative analysis framework. Neuroimage 145, 346–364 (2017).

35. Yang, Z. et al. A deep learning framework identifies dimensional representations of Alzheimer’s Disease from brain structure. Nat. Commun. 12, 7065 (2021).

36. Feng, Y. et al. Deep multiview learning to identify imaging-driven subtypes in mild cognitive impairment. BMC Bioinformatics 23, 402 (2022).

37. Sun, K. et al. Achieving multi-modal brain disease diagnosis performance using only single-modal images through generative AI. Commun Eng 3, (2024).

38. Meng, W. et al. Identifying sex-specific sub-phenotypes of Alzheimer’s disease progression using longitudinal electronic health records. EBioMedicine 130, 106391 (2026).

39. Peng, J., et al. Sparse MoE as a new treatment: Addressing forgetting, fitting, learning issues in multi-modal multi-task learning. (2023).

40. Shazeer, N., et al. Outrageously large neural networks: The Sparsely-Gated Mixture-of-experts layer. *arXiv [cs.LG]* (2017).

41. Han, X., Harris, C., Ho, N., Nguyen, H. & Saria, S. FuseMoE: Mixture-of-Experts Transformers for Fleximodal Fusion. in Advances in Neural Information Processing Systems 37 (eds. Globerson, A. et al.) vol. 37 67850–67900 (Neural Information Processing Systems Foundation, Inc. (NeurIPS), San Diego, California, USA, 2024).

42. Mustafa, B., Riquelme, C., Puigcerver, J., Jenatton, R. & Houlsby, N. Multimodal contrastive learning with LIMoE: The Language-image mixture of experts. arXiv [cs.CV] (2022).

43. Minoura, K., Abe, K., Nam, H., Nishikawa, H. & Shimamura, T. A mixture-of-experts deep generative model for integrated analysis of single-cell multiomics data. *Cell Rep*. Methods 1, 100071 (2021).

44. Ziquan, W., Tingting, D., Tianlong, C. & Guorong, W. BrainMoE: Cognition joint embedding via mixture-of-expert towards robust brain foundation model. https://openreview.net/pdf?id=05cVmYJJnb.

45. Xu, W., Jiang, H. & Liang, X. Leveraging knowledge of modality experts for incomplete multimodal learning. in Proceedings of the 32nd ACM International Conference on Multimedia 438–446 (ACM, New York, NY, USA, 2024).

46. DeepSeek-AI et al. DeepSeek-V3 Technical Report. *arXiv [cs.CL]* (2024).

47. Yun, S., et al. Flex-MoE: Modeling arbitrary modality combination via the Flexible Mixture-of-experts. in (Advances in Neural Information Processing Systems, 2024).

48. Dempster, A. P., Laird, N. M. & Rubin, D. B. Maximum likelihood from incomplete data via the *EM* algorithm. J. R. Stat. Soc. Series B Stat. Methodol. 39, 1–22 (1977).

49. Verdi, S. et al. Personalizing progressive changes to brain structure in Alzheimer’s disease using normative modeling. Alzheimers. Dement. 20, 6998–7012 (2024).

50. Wei, X. et al. Mapping heterogeneous brain structural subtypes in alzheimer’s disease and mild cognitive impairment using normative models. Transl. Psychiatry 16, 168 (2026).

51. Petersen, R. C. et al. Alzheimer’s Disease Neuroimaging Initiative (ADNI): clinical characterization. Neurology 74, 201–209 (2010).

52. Besser, L. et al. Version 3 of the national Alzheimer’s coordinating center’s Uniform Data Set. Alzheimer Dis. Assoc. Disord. 32, 351–358 (2018).

53. Sudlow, C. et al. UK biobank: an open access resource for identifying the causes of a wide range of complex diseases of middle and old age. PLoS Med. 12, e1001779 (2015).

54. Davies, D. L. & Bouldin, D. W. A Cluster Separation Measure. IEEE Trans. Pattern Anal. Mach. Intell. PAMI-1, 224–227 (1979).

55. Poldrack, R. A. Region of interest analysis for fMRI. Soc. Cogn. Affect. Neurosci. 2, 67–70 (2007).

56. Atkinson, A. C., Riani, M. & Cerioli, A. Principal Components Analysis. in Springer Series in Statistics 229–296 (Springer New York, New York, NY, 2004).

57. van der Maaten, L. & Hinton, G. Visualizing Data using t-SNE. J. Mach. Learn. Res. 9, 2579–2605 (2008).

58. McInnes, L., Healy, J. & Melville, J. UMAP: Uniform Manifold Approximation and Projection for Dimension Reduction. arXiv [stat.ML] (2018).

59. Euesden, J., Lewis, C. M. & O’Reilly, P. F. PRSice: Polygenic risk score software. Bioinformatics 31, 1466–1468 (2015).

60. Rosen, W. G., Mohs, R. C. & Davis, K. L. A new rating scale for Alzheimer’s disease. Am. J. Psychiatry 141, 1356–1364 (1984).

61. Petersen, R. C. et al. Mild cognitive impairment: clinical characterization and outcome. Arch. Neurol. 56, 303–308 (1999).

62. Hughes, C. P., Berg, L., Danziger, W. L., Coben, L. A. & Martin, R. L. A new clinical scale for the staging of dementia. Br. J. Psychiatry 140, 566–572 (1982).

63. Nasreddine, Z. S. et al. The Montreal Cognitive Assessment, MoCA: A brief screening tool for mild cognitive impairment. J. Am. Geriatr. Soc. 53, 695–699 (2005).

64. Weintraub, S. et al. Version 3 of the Alzheimer Disease Centers’ neuropsychological test battery in the Uniform Data Set (UDS). Alzheimer Dis. Assoc. Disord. 32, 10–17 (2018).

65. Tombaugh, T. N., Kozak, J. & Rees, L. Normative data stratified by age and education for two measures of verbal fluency: FAS and animal naming. Arch. Clin. Neuropsychol. 14, 167–177 (1999).

66. Reitan, R. M. Validity of the Trail Making Test as an organic impairment indicator. Perceptual and Motor Skills 8, 271–276 (1958).

67. Murray, M. E. et al. Neuropathologically defined subtypes of Alzheimer’s disease with distinct clinical characteristics: a retrospective study. Lancet Neurol. 10, 785–796 (2011).

68. de Vries, L. E., Huitinga, I., Kessels, H. W., Swaab, D. F. & Verhaagen, J. The concept of resilience to Alzheimer’s Disease: current definitions and cellular and molecular mechanisms. Mol. Neurodegener. 19, 33 (2024).

69. Corder, E. H. et al. Gene dose of apolipoprotein E type 4 allele and the risk of Alzheimer’s disease in late onset families. Science 261, 921–923 (1993).

70. Vagnoni, A. et al. Calsyntenin-1 mediates axonal transport of the amyloid precursor protein and regulates Aβ production. Hum. Mol. Genet. 21, 2845–2854 (2012).

71. Yan, X. et al. FRMD4A-cytohesin signaling modulates the cellular release of tau. J. Cell Sci. 129, 2003–2015 (2016).

72. Deming, Y. et al. The MS4A gene cluster is a key modulator of soluble TREM2 and Alzheimer’s disease risk. Sci. Transl. Med. 11, eaau2291 (2019).

73. Lambert, J. C. et al. Meta-analysis of 74,046 individuals identifies 11 new susceptibility loci for Alzheimer’s disease. Nat. Genet. 45, 1452–1458 (2013).

74. Kunkle, B. W. et al. Genetic meta-analysis of diagnosed Alzheimer’s disease identifies new risk loci and implicates Aβ, tau, immunity and lipid processing. Nat. Genet. 51, 414–430 (2019).

75. Picard, C. et al. Apolipoprotein B is a novel marker for early tau pathology in Alzheimer’s disease. Alzheimers. Dement. 18, 875–887 (2022).

76. Jansen, I. E. et al. Genome-wide meta-analysis identifies new loci and functional pathways influencing Alzheimer’s disease risk. Nat. Genet. 51, 404–413 (2019).

77. ADNI documentation – diagnostic. https://adni.loni.usc.edu/quick-start-guide-asset101625/diagnostic.html.

78. Longitudinal neurocognitive and clinical phenotype data - national Alzheimer’s coordinating center (NACC). *National Alzheimer’s Coordinating Center (NACC)* https://www.naccdata.org/about-nacc-data/longitudinal-neurocognitive-and-clinical-phenotype-data/.

79. Showcase homepage. https://biobank.ndph.ox.ac.uk/showcase/.

80. Pan, J. et al. MRISeqClassifier: A deep learning toolkit for precise MRI sequence classification. AMIA Summits Transl. Sci. Proc. 2025, 405–413 (2025).

81. Desikan, R. S. et al. An automated labeling system for subdividing the human cerebral cortex on MRI scans into gyral based regions of interest. Neuroimage 31, 968–980 (2006).

82. Cerezo, M. et al. The NHGRI-EBI GWAS Catalog: standards for reusability, sustainability and diversity. Nucleic Acids Res. 53, D998–D1005 (2025).

83. Danecek, P. et al. The variant call format and VCFtools. Bioinformatics 27, 2156–2158 (2011).

84. Chang, C. C. et al. Second-generation PLINK: rising to the challenge of larger and richer datasets. Gigascience 4, 7 (2015).

85. Miles, A., et al. scikit-allel: A Python package for exploring and analysing genetic variation data. https://zenodo.org/records/13772087 (2024).

86. Fortin, J.-P. et al. Harmonization of cortical thickness measurements across scanners and sites. Neuroimage 167, 104–120 (2018).

87. Barnes, J. et al. Head size, age and gender adjustment in MRI studies: a necessary nuisance? Neuroimage 53, 1244–1255 (2010).

88. O’Brien, L. M. et al. Statistical adjustments for brain size in volumetric neuroimaging studies: some practical implications in methods. Psychiatry Res. 193, 113–122 (2011).

89. Zhang, R., Sheng, J., Zhang, Q., Wang, J. & Wang, B. A review of multimodal fusion-based deep learning for Alzheimer’s disease. Neuroscience 576, 80–95 (2025).

90. Ferreira, D. et al. Distinct subtypes of Alzheimer’s disease based on patterns of brain atrophy: longitudinal trajectories and clinical applications. Sci. Rep. 7, (2017).

91. Kuhn, H. W. The Hungarian method for the assignment problem. Nav. Res. Logist. Q. 2, 83–97 (1955).

92. He, K., Zhang, X., Ren, S. & Sun, J. Deep residual learning for image recognition. in 2016 IEEE Conference on Computer Vision and Pattern Recognition (CVPR) 770–778 (IEEE, 2016).

93. Sheng, G., Wang, Q., Pei, C. & Gao, Q. Contrastive deep embedded clustering. Neurocomputing 514, 13–20 (2022).

94. Cai, J. et al. Efficient deep embedded subspace clustering. in 2022 IEEE/CVF Conference on Computer Vision and Pattern Recognition (CVPR) (IEEE, 2022). doi:10.1109/cvpr52688.2022.00012.

95. Macqueen, J. B. Some methods for classification and analysis of multivariate observations. in Proceedings of the Fifth Berkeley Symposium on Mathematical Statistics and Probability vol. 1 281–297 (University of California Press, Berkeley, 1967).

96. von Luxburg, U. A tutorial on spectral clustering. Stat. Comput. 17, 395–416 (2007).

97. Welch, B. L. The Generalization of ‘Student’s’ Problem when Several Different Population Variances are Involved. Biometrika 34, 28 (1947).

98. Liu, D. C. & Nocedal, J. On the limited memory BFGS method for large scale optimization. Math. Program. 45, 503–528 (1989).

99. Benjamini, Y. & Hochberg, Y. Controlling the false discovery rate: A practical and powerful approach to multiple testing. J. R. Stat. Soc. Series B Stat. Methodol. 57, 289–300 (1995).

100. Kaplan, E. L. & Meier, P. Nonparametric Estimation from Incomplete Observations. J. Am. Stat. Assoc. 53, 457 (1958).

