## Supplementary Information for "ADSMoE: A mixture-of-experts-based transformer autoencoder for discovering multimodal Alzheimer’s disease subtypes"

[1] Department of Health Outcomes & Biomedical Informatics, University of Florida, Gainesville, FL, 32610, USA
[2] Department of Pharmaceutics, University of Florida, Gainesville, FL, 32610, USA
[3] J. Crayton Pruitt Family Department of Biomedical Engineering, University of Florida, Gainesville, FL, 32610, USA
[4] Division of Computational Health Sciences, Department of Surgery, University of Minnesota, Minneapolis, MN, 55455, USA
[5] Department of Neurology and Fixel Institute for Neurological Disorders, University of Florida, Gainesville, FL, 32610, USA
[†] This study used data from the Alzheimer's Disease Neuroimaging Initiative (ADNI) database (adni.loni.usc.edu).
[*] **Correspondence author**:
Rui Yin, PhD
Department of Health Outcomes & Biomedical Informatics
University of Florida
Gainesville, Florida, USA


### Supplementary Figures

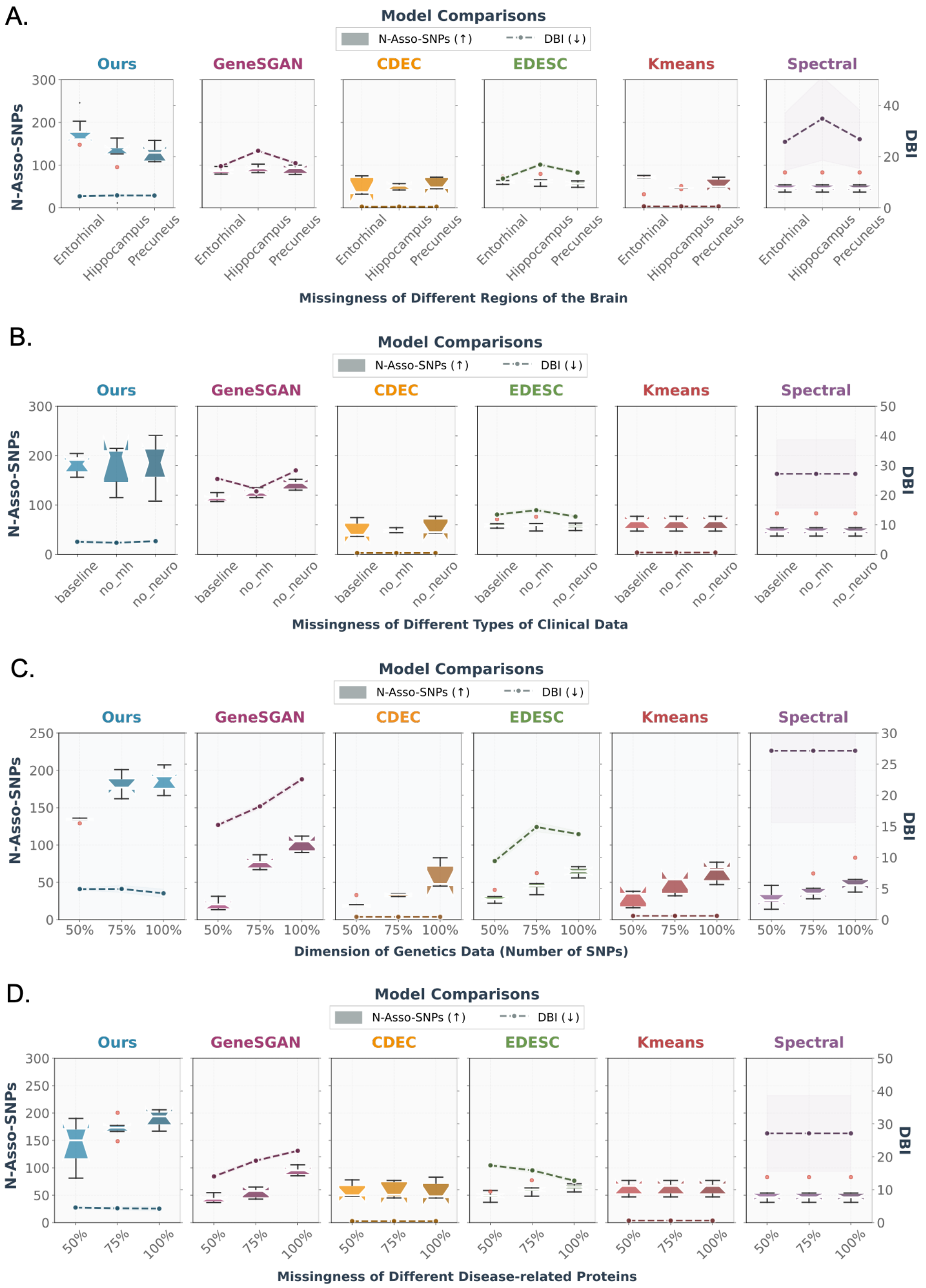


**Supplementary Figure S1. Robustness of ADSMoE under varying missingness and feature dimensions across multiple modalities. (A)** Comparison of subtyping performance under missingness of different brain regions (entorhinal cortex, hippocampus, and precuneus). Models are evaluated using the number of associated SNPs (N-Asso-SNPs; higher is better) and the Davies–Bouldin index (DBI; lower is better), demonstrating ADSMoE's robustness to region-specific imaging incompleteness. **(B)** Comparison of subtyping performance under missingness of different types of clinical data, including baseline (full clinical matrix), medical history, and neurological examination features. (**C**) Comparison of subtyping performance under varying dimensions of genetic features (50%, 75%, and 100%), illustrating the consistent performance of ADSMoE across genetic feature settings. (**D**) Comparison of subtyping performance under varying dimensions of protein features (50%, 75%, and 100%), illustrating the consistent performance of ADSMoE across protein feature settings.

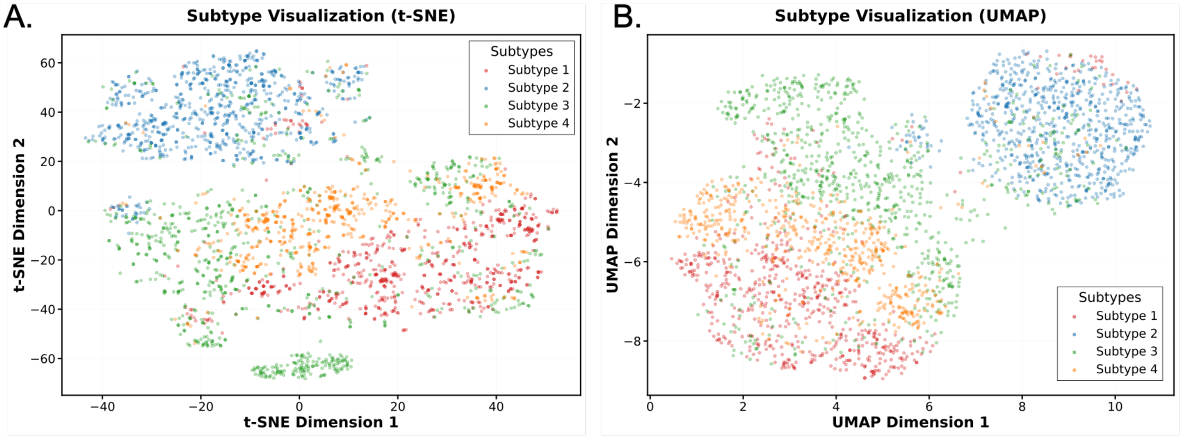


**Supplementary Figure S2. Visualization of ADSMoE-identified Alzheimer's disease subtypes (ADNI) in the latent space. (A)** Two-dimensional visualization of learned latent representations using t-distributed stochastic neighbor embedding (t-SNE). **(B)** Two-dimensional visualization of the same latent representations using uniform manifold approximation and projection (UMAP).

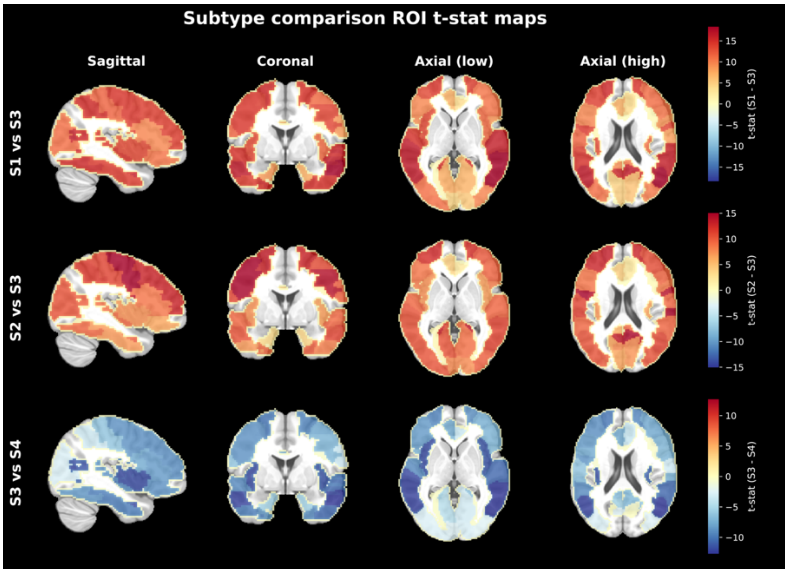


**Supplementary Figure S3. Pairwise comparison of subtype-specific structural MRI patterns using ROI-level t-statistic maps (ADNI).** Subtype-wise structural MRI differences are visualized by pairwise comparisons between subtypes, including S1 vs. S3, S2 vs. S3, and S3 vs. S4. For each comparison. Warmer colors indicate relatively larger regional brain volumes in the first subtype of each comparison, whereas cooler colors indicate relatively larger volumes in the second subtype. These maps highlight spatially distinct neuroanatomical patterns across AD subtypes.

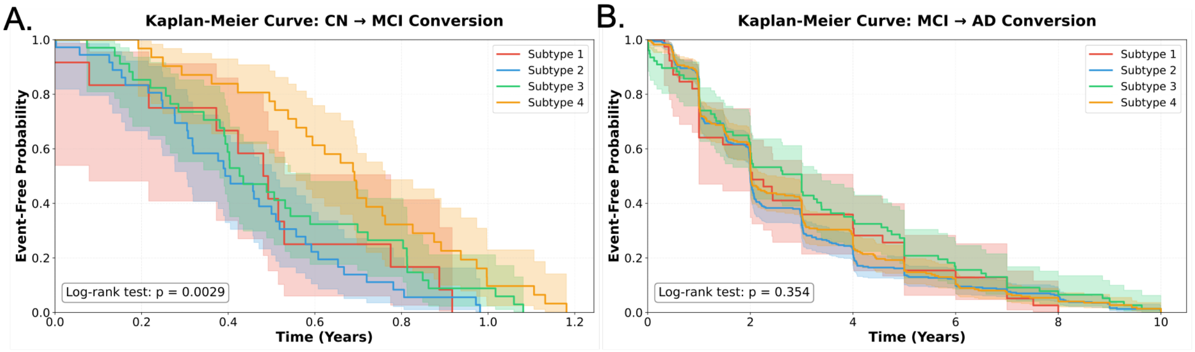


**Supplementary Figure S4. Retrospective comparison of diagnostic conversion times assessed by survival analysis (ADNI). (A)** Kaplan–Meier curves comparing conversion from cognitively normal (CN) to mild cognitive impairment (MCI) across the four identified subtypes. Event-free probability is shown over time, with shaded areas indicating confidence intervals. Differences among subtypes are evaluated using a log-rank test. Time to CN-to-MCI conversion differed significantly across subtypes (log-rank test, P = 0.0029). **(B)** Kaplan–Meier curves comparing conversion from MCI to Alzheimer's disease (AD) across subtypes. The difference among subtypes was not statistically significant (log-rank test, P = 0.354).

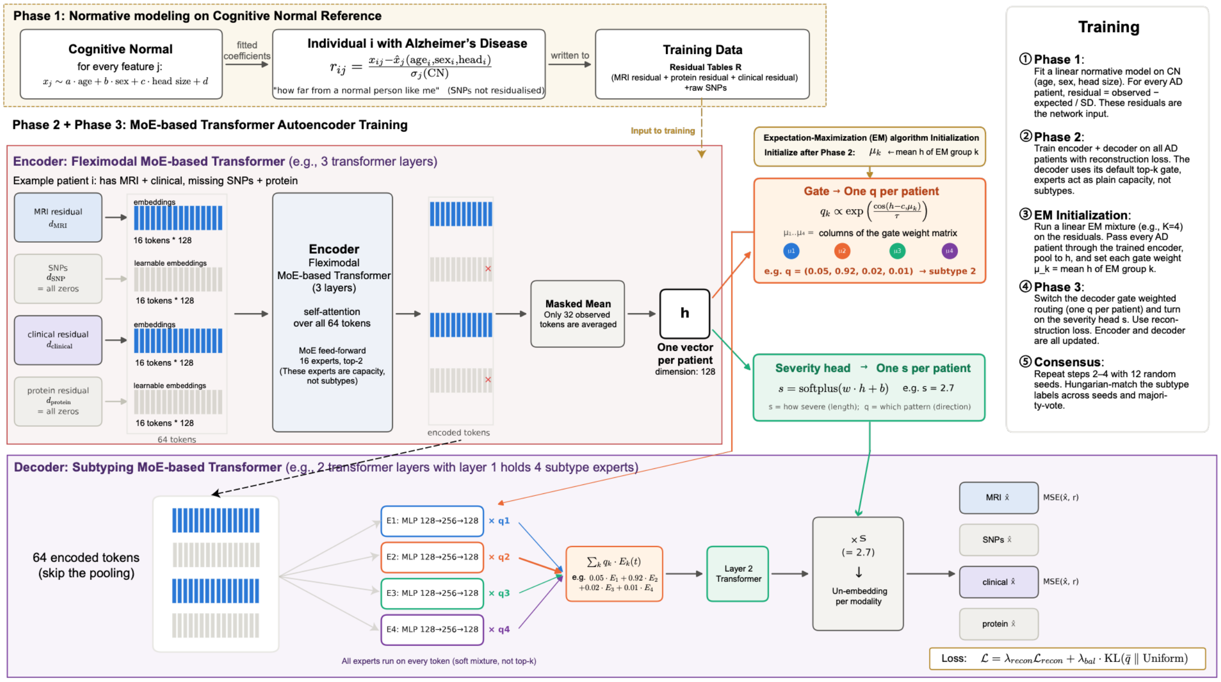


**Supplementary Figure S5. Detailed architecture and training workflow of ADSMoE.** Schematic illustration of the three-phase ADSMoE framework, including normative modeling using cognitively normal (CN) references, fleximodal MoE-based transformer autoencoder pretraining, and EM-initialized subtype-specific decoder training. The decoder employs patient-level soft routing across four subtype-specific experts and a separate severity head to reconstruct observed multimodal features, optimized using reconstruction and expert-balancing losses.

### Supplementary Tables

**Supplementary Table S1.** Baseline characteristics of participants (complete table of Table 1).

| **Cohort** | **ADNI** | | **NACC** | | **UKB** | |
|---|---|---|---|---|---|---|
| Variable | CN (N=1,159) | AD (N=876) | CN (N=759) | AD (N=876) | CN (N=876) | AD (N=876) |
| **Age** | | | | | | |
| <65 | 317 (27.4%) | 91 (10.4%) | 167 (22.0%) | 123 (14.0%) | 70 (8.0%) | 58 (6.6%) |
| 65-84 | 736 (63.5%) | 656 (74.9%) | 570 (75.1%) | 675 (77.1%) | 800 (91.3%) | 817 (93.3%) |
| >=85 | 45 (3.9%) | 58 (6.6%) | 22 (2.9%) | 78 (8.9%) | 0 (0.0%) | 1 (0.1%) |
| Missing | 61 (5.3%) | 71 (8.1%) | 0 (0.0%) | 0 (0.0%) | 6 (0.7%) | 0 (0.0%) |
| Age (Mean (SD)) | 71.5 (6.8) | 74.5 (7.4) | 71.6 (8.7) | 73.8 (8.5) | 71.9 (7.5) | 74.7 (6.2) |
| **Sex** | | | | | | |
| Male | 352 (30.4%) | 461 (52.6%) | 245 (32.3%) | 400 (45.7%) | 385 (43.9%) | 393 (44.9%) |
| Female | 479 (41.3%) | 344 (39.3%) | 514 (67.7%) | 476 (54.3%) | 491 (56.1%) | 483 (55.1%) |
| Unknown | 328 (28.3%) | 71 (8.1%) | 0 (0.0%) | 0 (0.0%) | 0 (0.0%) | 0 (0.0%) |
| **Race** | | | | | | |
| White | 691 (59.6%) | 748 (85.4%) | 552 (72.7%) | 688 (78.5%) | 830 (94.7%) | 850 (97.0%) |
| Black or African American | 91 (7.9%) | 33 (3.8%) | 151 (19.9%) | 144 (16.4%) | 12 (1.4%) | 13 (1.5%) |
| American Indian or Alaska Native | 2 (0.2%) | 0 (0.0%) | 15 (2.0%) | 8 (0.9%) | 0 (0.0%) | 0 (0.0%) |
| Native Hawaiian or Pacific Islander | 0 (0.0%) | 0 (0.0%) | 2 (0.3%) | 0 (0.0%) | 0 (0.0%) | 0 (0.0%) |
| Asian | 26 (2.2%) | 16 (1.8%) | 8 (1.1%) | 25 (2.9%) | 20 (2.3%) | 2 (0.2%) |
| Multiracial | 17 (1.5%) | 7 (0.8%) | 25 (3.3%) | 5 (0.6%) | 5 (0.6%) | 4 (0.5%) |
| Unknown or ambiguous | 332 (28.6%) | 72 (8.2%) | 6 (0.8%) | 6 (0.7%) | 9 (1.0%) | 7 (0.8%) |
| **Hispanic ethnicity** | | | | | | |
| No | 773 (66.7%) | 777 (88.7%) | 659 (86.8%) | 814 (92.9%) | N/A | |
| Yes | 53 (4.6%) | 24 (2.7%) | 95 (12.5%) | 59 (6.7%) | | |
| Missing/Unknown | 333 (28.7%) | 75 (8.6%) | 5 (0.7%) | 3 (0.3%) | | |
| **Year of education** | | | | | | |
| <=12 | 64 (5.5%) | 158 (18.0%) | 65 (8.6%) | 141 (16.1%) | 24 (2.7%) | 20 (2.3%) |

| | | | | | | |
|---|---|---|---|---|---|---|
| 13-16 | 357 (30.8%) | 373 (42.6%) | 336 (44.3%) | 382 (43.6%) | 471 (53.8%) | 483 (55.1%) |
| >=17 | 410 (35.4%) | 274 (31.3%) | 358 (47.2%) | 351 (40.1%) | 185 (21.1%) | 170 (19.4%) |
| Missing | 328 (28.3%) | 71 (8.1%) | 0 (0.0%) | 2 (0.2%) | 196 (22.4%) | 203 (23.2%) |
| Education (Mean (SD)) | 16.6 (2.5) | 15.6 (2.8) | 16.4 (4.4) | 16.0 (3.0) | 15.9 (3.4) | 15.4 (2.2) |
| **APOE Biomarkers** | | | | | | |
| No APOE ε4 allele | 516 (44.5%) | 268 (30.6%) | 480 (63.2%) | 355 (40.5%) | 590 (67.4%) | 281 (32.1%) |
| 1 copy of APOE ε4 allele | 203 (17.5%) | 376 (42.9%) | 135 (17.8%) | 256 (29.2%) | 250 (28.5%) | 380 (43.4%) |
| 2 copies of APOE ε4 allele | 19 (1.6%) | 133 (15.2%) | 6 (0.8%) | 68 (7.8%) | 21 (2.4%) | 196 (22.4%) |
| Missing or unknown or not assessed | 421 (36.3%) | 99 (11.3%) | 138 (18.2%) | 197 (22.5%) | 15 (1.7%) | 19 (2.2%) |
| **Modality Availability** | | | | | | |
| Image MRI | 632 (54.5%) | 848 (96.8%) | 759 (100.0%) | 876 (100.0%) | 80 (9.1%) | 61 (7.0%) |
| Protein Measurements | 490 (42.3%) | 329 (37.6%) | 0 (0.0%) | 0 (0.0%) | 572 (65.3%) | 647 (73.9%) |
| Genetics | 888 (76.6%) | 734 (83.8%) | 0 (0.0%) | 0 (0.0%) | 850 (97.0%) | 876 (100.0%) |
| Clinical | 928 (80.1%) | 740 (84.5%) | 759 (100.0%) | 876 (100.0%) | 876 (100.0%) | 876 (100.0%) |

“N/A” represents this variable and its related variables are not available in corresponding cohort.

**Supplementary Table S2.** Demographic characteristics of the four identified subtypes in ADNI.

| Variables | Subtype 1 (N=221) | Subtype 2 (N=227) | Subtype 3 (N=252) | Subtype 4 (N=176) |
|---|---|---|---|---|
| **Age** | | | | |
| <65 | 10 (4.5%) | 11 (4.8%) | 36 (14.3%) | 34 (19.3%) |
| 65-84 | 166 (75.1%) | 166 (73.1%) | 195 (77.4%) | 129 (73.3%) |
| >=85 | 19 (8.6%) | 24 (10.6%) | 9 (3.6%) | 6 (3.4%) |
| Missing | 26 (11.8%) | 26 (11.5%) | 12 (4.8%) | 7 (4.0%) |
| Age (Mean (SD)) | 76.3 (6.8) | 76.4 (6.8) | 73.4 (7.5) | 71.9 (7.5) |
| **Sex** | | | | |
| Male | 113 (51.1%) | 122 (53.7%) | 137 (54.4%) | 89 (50.6%) |
| Female | 82 (37.1%) | 79 (34.8%) | 103 (40.9%) | 80 (45.5%) |
| Missing | 26 (11.8%) | 26 (11.5%) | 12 (4.8%) | 7 (4.0%) |
| **Race** | | | | |
| White | 176 (79.6%) | 194 (85.5%) | 215 (85.3%) | 163 (92.6%) |
| Black | 11 (5.0%) | 4 (1.8%) | 16 (6.3%) | 2 (1.1%) |
| American Indian or Alaska Native | 0 (0.0%) | 0 (0.0%) | 0 (0.0%) | 0 (0.0%) |
| Asian | 7 (3.2%) | 2 (0.9%) | 6 (2.4%) | 1 (0.6%) |
| Multiracial | 1 (0.5%) | 1 (0.4%) | 3 (1.2%) | 2 (1.1%) |
| Unknown or ambiguous | 26 (11.8%) | 26 (11.5%) | 12 (4.8%) | 8 (4.5%) |
| **Hispanic ethnicity** | | | | |
| No | 190 (86.0%) | 195 (85.9%) | 233 (92.5%) | 159 (90.3%) |
| Yes | 4 (1.8%) | 6 (2.6%) | 5 (2.0%) | 9 (5.1%) |
| Missing/Unknown | 27 (12.2%) | 26 (11.5%) | 14 (5.6%) | 8 (4.5%) |
| **Year of education** | | | | |
| <=12 | 52 (23.5%) | 26 (11.5%) | 35 (13.9%) | 45 (25.6%) |
| 13-16 | 51 (23.1%) | 123 (54.2%) | 125 (49.6%) | 74 (42.0%) |
| >=17 | 92 (41.6%) | 52 (22.9%) | 80 (31.7%) | 50 (28.4%) |
| Missing | 26 (11.8%) | 26 (11.5%) | 12 (4.8%) | 7 (4.0%) |
| Education (Mean (SD)) | 15.7 (3.2) | 15.5 (2.7) | 15.9 (2.6) | 15.1 (2.8) |

**Supplementary Table S3.** Demographic characteristics of the four identified subtypes in NACC.

| Variables | Subtype 1 (N=445) | Subtype 2 (N=284) | Subtype 3 (N=28) | Subtype 4 (N=119) |
|---|---|---|---|---|
| **Age** | | | | |
| <65 | 43 (9.7%) | 34 (12.0%) | 8 (28.6%) | 38 (31.9%) |
| 65-84 | 356 (80.0%) | 229 (80.6%) | 18 (64.3%) | 72 (60.5%) |
| >=85 | 46 (10.3%) | 21 (7.4%) | 2 (7.1%) | 9 (7.6%) |
| Missing | 0 (0.0%) | 0 (0.0%) | 0 (0.0%) | 0 (0.0%) |
| Age (Mean (SD)) | 74.8 (8.0) | 74.0 (7.9) | 71.1 (10.9) | 70.2 (10.0) |
| **Sex** | | | | |
| Male | 197 (44.3%) | 135 (47.5%) | 12 (42.9%) | 56 (47.1%) |
| Female | 248 (55.7%) | 149 (52.5%) | 16 (57.1%) | 63 (52.9%) |
| Missing | 0 (0.0%) | 0 (0.0%) | 0 (0.0%) | 0 (0.0%) |
| **Race** | | | | |
| White | 313 (70.3%) | 241 (84.9%) | 23 (82.1%) | 111 (93.3%) |
| Black | 100 (22.5%) | 33 (11.6%) | 5 (17.9%) | 6 (5.0%) |
| American Indian or Alaska Native | 6 (1.3%) | 2 (0.7%) | 0 (0.0%) | 0 (0.0%) |
| Asian | 20 (4.5%) | 4 (1.4%) | 0 (0.0%) | 1 (0.8%) |
| Multiracial | 3 (0.7%) | 1 (0.4%) | 0 (0.0%) | 1 (0.8%) |
| Unknown or ambiguous | 3 (0.7%) | 3 (1.1%) | 0 (0.0%) | 0 (0.0%) |
| **Hispanic ethnicity** | | | | |
| No | 413 (92.8%) | 265 (93.3%) | 26 (92.9%) | 110 (92.4%) |
| Yes | 32 (7.2%) | 16 (5.6%) | 2 (7.1%) | 9 (7.6%) |
| Missing/Unknown | 0 (0.0%) | 3 (1.1%) | 0 (0.0%) | 0 (0.0%) |
| **Year of education** | | | | |
| <=12 | 101 (22.7%) | 7 (2.5%) | 6 (21.4%) | 27 (22.7%) |
| 13-16 | 114 (25.6%) | 214 (75.4%) | 16 (57.1%) | 38 (31.9%) |
| >=17 | 229 (51.5%) | 62 (21.8%) | 6 (21.4%) | 54 (45.4%) |
| Missing | 1 (0.2%) | 1 (0.4%) | 0 (0.0%) | 0 (0.0%) |
| Education (Mean (SD)) | 16.0 (3.6) | 16.1 (2.1) | 15.6 (2.7) | 15.9 (2.8) |

**Supplementary Table S4.** Demographic characteristics of the four identified subtypes in UKB.

| Variables | Subtype 1 (N=275) | Subtype 2 (N=252) | Subtype 3 (N=258) | Subtype 4 (N=91) |
|---|---|---|---|---|
| **Age** | | | | |
| <65 | 18 (6.5%) | 22 (8.7%) | 16 (6.2%) | 2 (2.2%) |
| 65-84 | 257 (93.5%) | 229 (90.9%) | 242 (93.8%) | 89 (97.8%) |
| >=85 | 0 (0.0%) | 1 (0.4%) | 0 (0.0%) | 0 (0.0%) |
| Age (Mean (SD)) | 74.7 (6.3) | 74.0 (6.6) | 75.0 (6.1) | 75.6 (4.9) |
| **Sex** | | | | |
| Male | 134 (48.7%) | 98 (38.9%) | 121 (46.9%) | 40 (44.0%) |
| Female | 141 (51.3%) | 154 (61.1%) | 137 (53.1%) | 51 (56.0%) |
| Missing | 0 (0.0%) | 0 (0.0%) | 0 (0.0%) | 0 (0.0%) |
| **Race** | | | | |
| White | 268 (97.5%) | 240 (95.2%) | 254 (98.4%) | 88 (96.7%) |
| Black | 3 (1.1%) | 6 (2.4%) | 4 (1.6%) | 0 (0.0%) |
| American Indian or Alaska Native | 0 (0.0%) | 0 (0.0%) | 0 (0.0%) | 0 (0.0%) |
| Asian | 2 (0.7%) | 0 (0.0%) | 0 (0.0%) | 0 (0.0%) |
| Multiracial | 2 (0.7%) | 1 (0.4%) | 0 (0.0%) | 1 (1.1%) |
| Unknown or ambiguous | 0 (0.0%) | 5 (2.0%) | 0 (0.0%) | 2 (2.2%) |
| **Year of education** | | | | |
| <=12 | 4 (1.5%) | 9 (3.6%) | 6 (2.3%) | 1 (1.1%) |
| 13-16 | 144 (52.4%) | 141 (56.0%) | 144 (55.8%) | 54 (59.3%) |
| >=17 | 57 (20.7%) | 44 (17.5%) | 51 (19.8%) | 18 (19.8%) |
| Missing | 70 (25.5%) | 58 (23.0%) | 57 (22.1%) | 18 (19.8%) |
| Education (Mean (SD)) | 15.3 (2.3) | 15.4 (1.8) | 15.2 (2.4) | 16.4 (2.0) |

**Supplementary Table S5.** Settings of the compared models.

| Model | Features | Hyperparameters | link |
|---|---|---|---|
| ADSMoE | Image + Genetics + Protein + Clinical | Encoder expert number = 16, Decoder expert number = 4, Encoder top k = 2, Decoder routing = dense soft mixture, Hidden dimension = 128, Encoder transformer layers = 3, Decoder transformer layers = 2, Batch size = 32, Learning rate = $1\times10^{-4}$, Encoder training epochs = 40, Joint training epochs = 30, EM initialization K = 4, reconstruction loss = 1.0, expert balancing loss = 0.01. | See code availability |
| GeneSGAN | Image + Genetics (Protein + Clinical as covariates) | Max epochs = 30000, Batch size = 8, Generator learning rate = 0.0002, Wasserstein distance weight = 0.18, Adversarial quotient = 35, Cluster loss weight = 0.03, Reconstruction weight (λ) = 15, Mutual information weight (μ) = 5, Lipschitz constant = 0.5 | https://github.com/zhijian-yang/GeneSGAN |
| CDEC | Image + Genetics + Protein + Clinical | Encoder layers = [500, 500, 2000, 10], Decoder layers = [10, 2000, 500, 500], Latent dimension = 10, Training epochs = 200, Batch size = 8, Learning rate = 0.001, Contrastive loss weight (α) = 0.05, post-clustering = K-Means (K=4), Optimizer = Adam | https://github.com/guoshuaiS/contrastive-deep |
| EDESC | Image + Genetics + Protein + Clinical | Encoder layers = [500, 500, 1000, 40], Decoder layers = [40, 1000, 500, 500], Latent dimension = 40, Subspace dimension = 10, Pretrain epochs = 50, Training epochs = 200, Learning rate = 0.001, Self-expressive coefficient (η) = 5, Balance weight (β) = 0.1, Optimizer = Adam | https://github.com/JinyuCai95/EDESC-pytorch |
| KMeans | Image + Genetics + Protein + Clinical | Initialization = k-means++, n_init = 200, max_iter = 500 | scikit-learn==0.24.2 |
| Spectral clustering | Image + Genetics + Protein + Clinical | Affinity = nearest_neighbors kernel, n_init = 200 | scikit-learn==0.24.2 |

### Description of Additional Supplementary Files

Supplementary Data contains five worksheets with feature annotations and selected numerical results for Figures 3F, 4C, 4D, and 5A. The first table lists the region-of-interest (ROI) indices used in the imaging analyses, including ROI identifiers, anatomical region names, and corresponding brain region groupings, which support the ROI-level z-score patterns shown in **Figure 3F**. The second table reports full results of SNP–subtype association analyses, including likelihood ratio test statistics, p-values, and false discovery rate (FDR)–adjusted significance levels, corresponding to the genetic association results shown in **Figure 4C**. The third table summarizes subtype-associated protein analyses and presents statistical results for all proteins with significant differences across subtypes after multiple-testing correction, as shown in **Figure 4D**. The fourth table lists the code meanings for clinical variables shown in **Figure 5A**. The fifth table provides detailed DIABLO component-based similarity scores among genetic variants, imaging-derived regional measures, clinical variables, and protein features, which form the basis of the Circos diagram visualization in **Figure 5A**.
